# The Utility of Long-Read Sequencing in Diagnosing Genetic Autosomal Recessive Parkinson’s Disease: a genetic screening study

**DOI:** 10.1101/2024.06.14.24308784

**Authors:** Kensuke Daida, Hiroyo Yoshino, Laksh Malik, Breeana Baker, Mayu Ishiguro, Rylee Genner, Kimberly Paquette, Yuanzhe Li, Kenya Nishioka, Satoshi Masuzugawa, Makito Hirano, Kenta Takahashi, Mikhail Kolmogolv, Kimberley J Billingsley, Manabu Funayama, Cornelis Blauwendraat, Nobutaka Hattori

## Abstract

**Background:** Mutations within the genes *PRKN* and *PINK1* are the leading cause of early onset autosomal recessive Parkinson’s disease (PD). However, the genetic cause of most early-onset PD (EOPD) cases still remains unresolved. Long-read sequencing has successfully identified many pathogenic structural variants that cause disease, but this technology has not been widely applied to PD. We recently identified the genetic cause of EOPD in a pair of monozygotic twins by uncovering a complex structural variant that spans over 7 Mb, utilizing Oxford Nanopore Technologies (ONT) long-read sequencing. In this study, we aimed to expand on this and assess whether a second variant could be detected with ONT long-read sequencing in other unresolved EOPD cases reported to carry one heterozygous variant in *PRKN* or *PINK1*.

**Methods:** ONT long-read sequencing was performed on patients with one reported *PRKN/PINK1* pathogenic variant. EOPD patients with an age at onset younger than 50 were included in this study. As a positive control, we also included EOPD patients who had already been identified to carry two known *PRKN* pathogenic variants. Initial genetic testing was performed using either short-read targeted panel sequencing for single nucleotide variants and multiplex ligation-dependent probe amplification (MLPA) for copy number variants.

**Results:** 48 patients were included in this study (*PRKN* “one-variant” n = 24, *PINK1* “one-variant” n = 12, *PRKN* “two-variants” n = 12). Using ONT long-read sequencing, we detected a second pathogenic variant in six *PRKN* “one-variant” patients (26%, 6/23) but none in the *PINK1* “one-variant” patients (0%, 0/12). Long-read sequencing identified one case with a complex inversion, two instances of structural variant overlap, and three cases of duplication. In addition, in the positive control *PRKN* “two-variants” group, we were able to identify both pathogenic variants in *PRKN* in all the patients (100%, 12/12).

**Conclusions:** This data highlights that ONT long-read sequencing is a powerful tool to identify a pathogenic structural variant at the *PRKN* locus that is often missed by conventional methods. Therefore, for cases where conventional methods fail to detect a second variant for EOPD, long-read sequencing should be considered as an alternative and complementary approach.

## Background

Parkinson’s disease (PD) is a neurodegenerative disorder showing motor symptoms, including resting tremor, rigidity, bradykinesia, and postural instability. These motor symptoms are caused by loss of dopaminergic neurons in the substantia nigra pars compacta. PD is considered to be caused by a combination of genetics, environment, and aging.[1] According to genome-wide association studies (GWAS), the heritability of PD is estimated to be around 16 - 36 %.[2] But taken together, the known GWAS PD risk loci can only explain part of this heritability, meaning that most of the common genetic variation contributing to disease risk is still unknown.

Approximately 5-10% of all PD cases can be attributed to a “monogenic” cause of disease. Biallelic *PRKN* and *PINK1* mutations are known to be the frequent cause of early onset PD (EOPD) and autosomal recessive PD.[3,4] The frequency of *PRKN* mutations increases with lower age at onset (AAO) of PD and is estimated to account for 77% in the PD patients with AAO younger than 20.[5] Typically, biallelic *PRKN* and *PINK1* variant carriers are characterized by early onset Parkinsonism, foot dystonia, sleep benefit, and good response to levodopa.

Intriguingly, monoallelic variants (carriers of 1 damaging variant) of *PRKN* and *PINK1* have been considered to be associated with PD.[6–9] Monoallelic *PRKN* or *PINK1* carriers are estimated to account for 2% of all PD patients.[10] However, some studies showed negative association with PD and heterozygous variants of *PRKN* and *PINK1*.[11–13] Therefore, the role of monoallelic *PRKN/PINK1* variants remains controversial.

*PRKN* and *PINK1* can harbor pathogenic variants that include single nucleotide variants (SNVs), exon dosage variations, and complex rearrangements.[14] Using long-read sequencing, we recently identified a heterozygous large inversion of *PRKN*, which was missed by multiplex ligation-dependent probe amplification (MLPA) and short-read targeted resequencing, in monozygotic twins with EOPD known to have a heterozygous exon 3 deletion.[15] Expanding on this, using long-read sequencing we assessed how often complex structural variants (SVs), like inversions, are missed by short read sequencing and MLPA in young-onset PD patients who only carry monoallelic *PRKN* and *PINK1* variants.

## Methods

### Study design and participants

All the participants were selected according to following criteria; 1) AAO of PD is younger than the age of 50, 2) the participant was confirmed to have one pathogenic variant in *PRKN* or *PINK1* based on targeted resequencing of PD related genes and MLPA, 3) the participant does not have any pathogenic variants in other known PD or dementia related genes (*SNCA, UCHL1, DJ-1, ATP13A2, GIGYF2, HTRA2, PLA2G6, FBXO7, VPS35, EIF4G1, DNAJC6, SYNJ1, DNAJC13, CHCHD2, GCH1, NR4A2, VPS13C, RAB7L1, BST1, C19orf12, RAB39B, MAPT, PSEN1, GRN, APP,* and *APOE)*. We also included patients with two known *PRKN* variants to assess the overall performance of long-read sequencing to detect these variants.

All the participants underwent a neurological examination and clinical information was collected by the attending neurologist. PD was clinically diagnosed according to standard clinical criteria.[16,17] DNA was extracted from peripheral blood by the standard protocol using QIAamp DNA Blood Maxi Kit (QIAGEN, Venlo, Netherlands). Study design is visualized in Figure 1.

**Figure 1.**
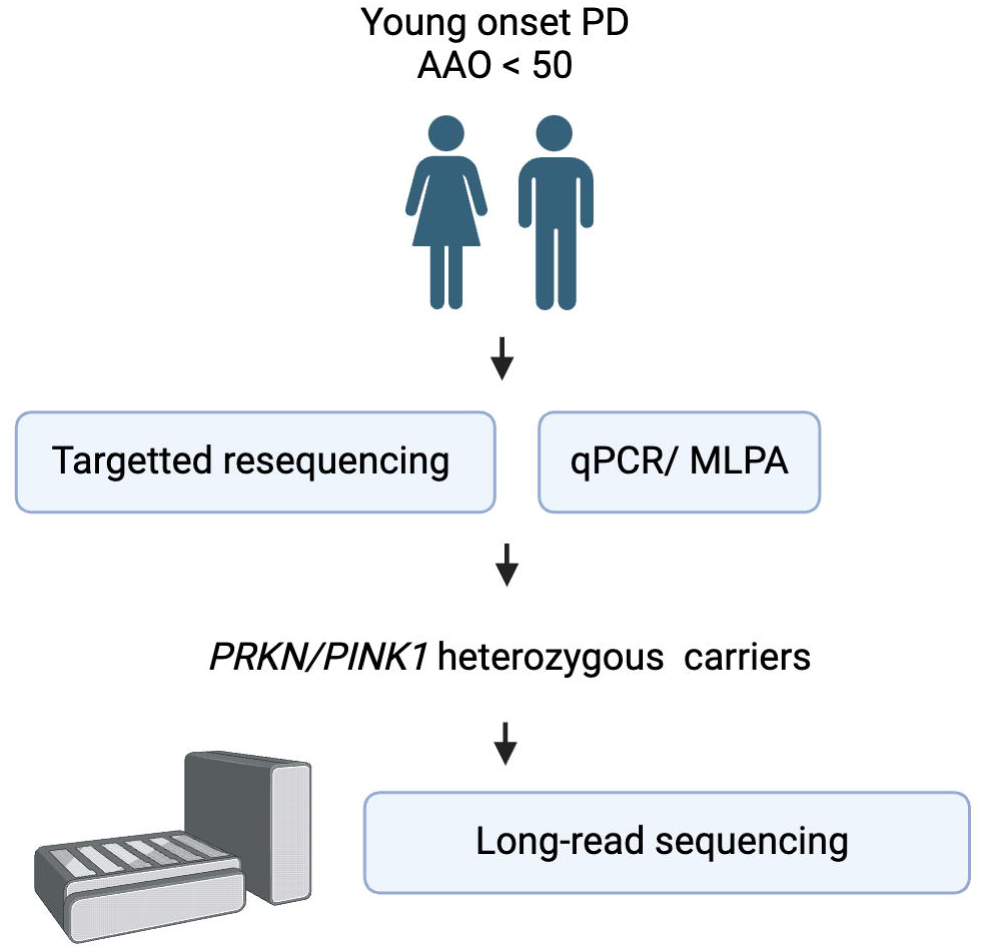
Analysis Workflow Schematic. PD; Parkinson’s disease, AAO; Age of onset, MLPA; Multiplex ligation-dependent probe amplification

### Genetic Testing

#### Targeted panel sequencing using short read sequencing

The targeted panel sequencing was performed to sequence for PD-related genes has been previously reported.[9] In brief, Ion Torrent system (Thermo Fisher Scientific, Waltham, MA, USA) was used for sequencing and we selected rare variants with an allelic frequency under 0.001 for autosomal dominant inheritance and under 0.005 for autosomal recessive inheritance by referring to public gene databases and annotating them using several prediction tools to define the pathogenicity of the variant.

#### Structural variants screening

Initially, copy number variants (CNVs) of *PRKN* were analyzed by quantitative PCR (qPCR) with TaqMan probe (Applied Biosystems, Foster City, CA, USA) using ABI PRISM 7700 sequence detection system (Applied Biosystems, Foster City, CA, USA) or by multiplex MLPA using SALSA MLPA Probemix P051 Parkinson mix (MRC-Holland, Amsterdam, the Netherlands), as we reported previously.[9,18] Secondly, for cases where we found either one variant of *PRKN* or *PINK1* after targeted resequencing and the initial qPCR/MLPA analysis, we conducted a second MLPA experiment. This involved using the MLPA P052 Parkinson probe mix (MRC-Holland, Amsterdam, Netherlands) to standardize the method for screening CNVs. We determined the number of *PRKN*/*PINK1* CNVs by considering the results from both the initial (qPCR/MLPA with P051) and the second (MLPA with P052) screenings. MLPA procedures were performed according to the manufacturer’s instructions.

#### Oxford Nanopore Technologies long-read sequencing

We used the DNA prepared for the short-read sequencing for the long-read sequencing. Sequencing was prepared according to our protocol reported previously.[19],[20]. In brief, DNA samples were sized using the Femto Pulse (Agilent Technologies Santa Clara, CA, USA) and run on the Sage BluePippin system (Sage science Beverly, MA, USA) to remove DNA fragments below 10kb. Libraries were prepared using the Kit V14 Ligation sequencing kit from Oxford Nanopore Technologies (ONT) and sequenced using PromethION for 72 hours on a R10.4.1 flow cell (Oxford Nanopore Technologies, Oxford, UK). Base calling was performed by Dorado v0.3.4 (https://github.com/nanoporetech/dorado) and Minimap v2.26 was used to map the reads to the GRCh38 reference genome. Sniffles v2.2 and CuteSV v2.0.3, and Seversus v0.1.2 were used for calling SVs.[21–23] SVs were annotated by AnnotSV v3.1.1.[24] All the identified variants in at least one SV caller were confirmed visually by integrative genome viewer (IGV).[25] SNVs were called by Clair3 and the output vcf was annotated using Annovar.[26,27] To phase the variants, PEPPER-Margin-Deep-Variant v0.8 was used with -phased_output option.[28] Additional adaptive long-read sequencing for *PRKN* and *PINK1* was performed in 12 samples to increase the coverage of *PRKN* and *PINK1* using ONT recommend guidelines.

#### Confirmation of SVs

To confirm the complex SVs that were only identified by long-read sequencing, we amplified the breakpoints region using PCR and performed Sanger sequencing by the primers specifically designed by Primer 3 (eTable 1).

#### Statistics

We compared the clinical phenotype between *PRKN* one-variant carriers and *PRKN* two variants carriers after ONT long-read sequencing by Pearson’s correlation coefficient and point-biserial correlation coefficient.

## Results

### Data overview

We included 48 patient samples (*PRKN* one-variant group n = 24, *PRKN* two-variants group n = 12, *PINK1* one-variant group n = 12; Female : Male = 28 : 20) in this study (Table 1). DNA quality control results for long-read sequencing identified one sample that did not meet criteria for inclusion, so a total of 47 samples were included in the analysis (eTable 2-4). The overall data output for long-read genome sequencing is 98.5 ± 25.04 Gb, and N50 was 20.3 ± 2.82 Kb for genome sequencing, and for adaptive sampling the data output is 13.1 ± 7.17 Gb, and N50 was 1.60 ± 1.68 Kb (eTable 5-6, eFigure 1).

**Table 1.** Study population.

|  | ***PRKN* hetero** | ***PINK1* hetero** | ***PRKN* homo** |
| --- | --- | --- | --- |
|  | *one-variant group* | *one-variant group* | *two-variants group* |
| N | 24 | 12 | 12 |
| Age at onset | 37.4 ± 7.86 | 32.7 ± 9.70 | 27.7 ± 10.06 |
| Age at examination | 50.0 ± 12.19 | 46.3 ± 13.55 | 47.4 ± 13.68 |
| Female : Male | 16 : 8 | 5:7 | 7:5 |
| Known Heterozygous variant (SNV/SV) | 10/14 | 12/0 | 4/20 |

SNV; single nucleotide variant, SV; structural variant, hetero; heterozygous, homo; homozygous

### Assessing the performance of long-read sequencing in *PRKN*-PD

To assess the overall performance of long-read sequencing, we included 12 *PRKN* patient samples carrying two known *PRKN* variants. All the known variants including SNVs and SVs identified by panel sequencing, qPCR, and MLPA were identified and confirmed using long-read sequencing (Table 2), showing 100% accuracy. In one case (PRKN-18), in which duplication of exon 2 and deletion of exon 5 were found by MLPA, long-read sequencing was able to more accurately define the genomic events and reported that there was a duplication of exon 2 to 4 and deletion of exon 3 to 5. In three cases (PRKN-12, PRKN-24, PRKN-27), SVs were identified as duplications by MLPA but were categorized as inversions by long-read sequencing SV callers. Visual inspection using the IGV indicated that one allele of the duplication was inverted, similarly showing the value of MLPA but also the superiority of long-read sequencing in defining what the actual variant is.

**Table 2.** Comparison of long-read and conventional sequencing methods.

|  | one variants of *PRKN* in *MLPA* and targeted resequencing | two variants of *PRKN* in *MLPA* and targeted resequencing | one variants of *PINK1* in *MLPA* and targeted resequencing |
| --- | --- | --- | --- |
| N | 23 | 12 | 12 |
| Two variants carriers by long-read seq | 26.1 % (6/23) | 100 % (12 /12) | 0% (0/12) |
| LRS miss | 0 | 0 | 0 |
| discordant with LRS and MLPA | 8<sup>a</sup> | 1<sup>b</sup> | 0 |

a) Including six newly identified two variant carriers, one with complex variant (DUP-NML-DUP/INV), and one with size difference of deletion between long-read sequencing and MLPA.
b) One sample has an overlapping copy number variant on different alleles which makes the size of structural variants different between long-read sequencing and MLPA.
LRS; long-read sequencing, MLPA; multiplex ligation-dependent probe amplification, DUP; duplication, NML; normal, INV; inversion

### Identification of the second *PRKN* variant in patients with single *PRKN* variant

In the other 35 patients with one *PRKN* or *PINK1* variant, long-read sequencing identified a second, previously undetected pathogenic variant in 6 out of 23 patients with one variant in the *PRKN* gene (26.1%), whereas no additional variants were found in the 12 patients with a heterozygous variant in the *PINK1* gene (0%) (Table 2). Notably, in the case of PRKN-31, long-read sequencing with one SV caller (Severus) detected a deletion in exon 3, a finding that was not reported by the other two SV callers, Sniffles and CuteSV. However, manual inspection using IGV revealed that this was actually a deletion spanning exons 3 and 4, underscoring the importance of manual curation (eFigure 2).

In the six “new” two-variants *PRKN* cases identified only by long-read sequencing, one case had inversion, three cases showed overlapping pathogenic *PRKN* SVs in each allele, and two cases were carrying a duplication (Table 3). Specifically, a complex inversion including exon 3 was identified from a patient (PRKN-10) previously recognized to have an exon 3 deletion via MLPA (Figure 2). All SV callers identified two overlapping inversions in the same region including exon 3. Detailed analysis using the IGV by linking mapped reads revealed an inversion involving the region of exon 3, along with duplication of the flanking regions on both sides of exon 3 (Figure 2A,2B). Additionally, we also confirmed the known exon 3 deletion in another allele by IGV. To confirm the inversion and deletion, we amplified the sequence of breakpoints and performed Sanger sequencing to ensure that the sequences surrounding the breakpoints were the same for long reads and Sanger sequencing (Figure 2C). This variant appeared to be an inversion of region including exon 3 accompanied by the duplications of flanking regions.

**Figure 2.**
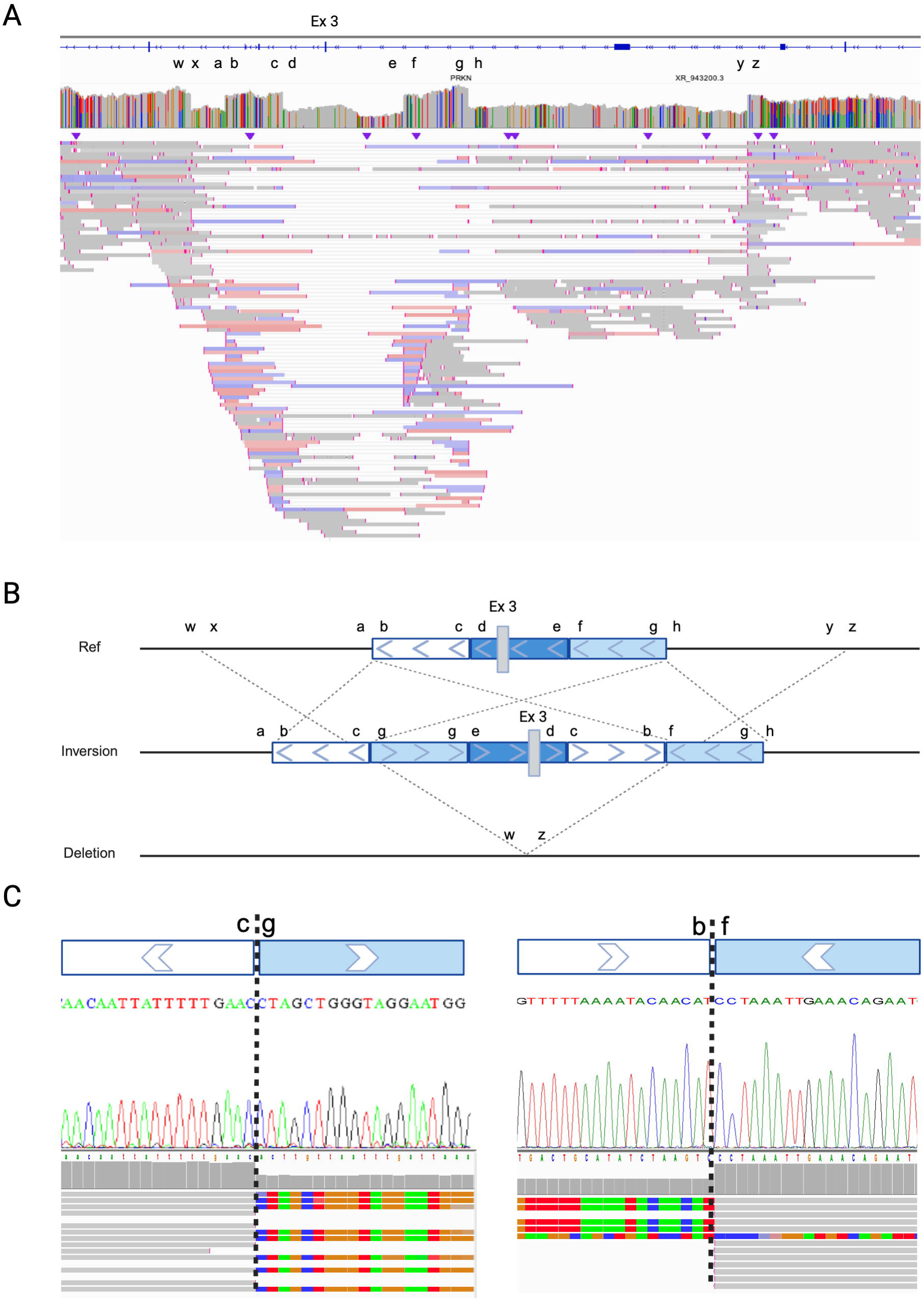
Description for complex inversion including exon 3 of *PRKN*. (A) Screenshot from Integrative Genomics Viewer (IGV) showcasing the complex inversion containing exon 3 and deletion of exon 3 in another allele. Notably, segments a-h and w-z within this diagram are directly aligned with corresponding segments in panels (B) and (C). (B) A schematic illustration depicting genetic variations: the middle section outlines a complex inversion, while the lower section details a deletion, both in comparison to the reference sequence shown in the upper section. Notably, the region encompassing exon 3 undergoes inversion within segments b-g, with the surrounding areas of exon 3 (b-c and f-g) being duplicated. (C) Results from Sanger sequencing, focusing on the inversion breakpoints. IGV; Integrative Genomics Viewer, INV; inversion, DUP;duplication, NML; normal

**Table 3.** Samples with two *PRKN* variants identified only by Long-read sequencing.

|  | **Variants by MLPA and short-read** | **Variant one by long-read seq** | **Variant two by long-read seq** | **Notes** |
| --- | --- | --- | --- | --- |
| PRKN-10 | *PRKN* exon 3 deletion | *PRKN* exon 3 deletion | *PRKN* exon 3 complex inversion | complex structural variant |
| PRKN-11 | *PRKN* exon 2 deletion | *PRKN* exon 2 deletion & exon 3-4 duplication | *PRKN* exon 4 deletion | deletion and duplication overlap |
| PRKN-21 | *PRKN* c.535-3A>G (T>C) (p.G179RfsX10) | *PRKN* c.535-3A>G(p.G179RfsX10) | *PRKN* exon 6 duplication | duplication |
| PRKN-25 | *PRKN* exon 5 - 6 duplication | *PRKN* exon 6 duplication | *PRKN* exon 5-6 duplication | duplication overlap |
| PRKN-32 | *PRKN* exon 2-3 deletion | *PRKN* exon 3 deletion | *PRKN* exon 2 deletion | deletion continuous |
| PRKN-35 | *PRKN* c.536delG_p.G179Vfs*9 | *PRKN* c.536delG(p.G179Vfs*9) | *PRKN* exon 6 duplication | duplication |

MLPA; multiplex ligation-dependent probe amplification

Three subjects (PRKN-11, PRKN-25, PRKN-32) presented with overlap of duplication and/or deletion in the same allelic exons which makes it difficult to identify and judge by MLPA. PRKN-11, harboring exon 2 deletion identified by MLPA, was identified to have duplication of exon 3-4, exon 2 deletion, and exon 4 deletion (eFigure 2). PEPPER-Margin-DeepVariant was not able to phase the variants around these exons likely due to insufficient sequence length (N50). However, manual phasing determined that the duplication of exon 3-4 and exon 2 deletion were located on the same allele, while the exon 4 deletion was on another allele. PRKN-25 was identified to have exon 6 duplication and exon 5-6 duplication by long-read sequencing. In MLPA, since the two duplications overlapped, it is not possible to differentiate those two variants (eFigure 2). PRKN-32, who was identified to have exon 2-3 deletion by MLPA, appeared to have separate deletions of exon 2 and exon 3 deletion (eFigure 2). The absence of a heterozygous variant between these deletions made phasing challenging, but the patient’s phenotype suggests that the two variants are unlikely to be on the same allele. Additionally, two cases (PRKN-21 and PRKN-35) carried duplications which were not detected by MLPA (eFigure 2).

Alongside these duplications, PRKN-21 and PRKN-35 carried known pathogenic SNVs (c.535-3A>G (p.G179RfsX10) and c.536delG(p.G179Vfs*9)) that were identified by short-read targeted sequencing which were also confirmed by long-read sequencing.

One case, PRKN-9, was characterized by a complex SV labeled as DUP-NML-INV/DUP.[29] MLPA indicated that this patient carried an exon 7 multiplication. However, long-read sequencing identified not only an exon 7 multiplication but also an apparent increase of sequence reads overlapping with exon 6 and 7, suggesting the presence of an additional duplication (Figure 3). Further examination of split reads revealed a unique pattern. The reads mapped at two distinct breakpoints (eFigure 3. A-B and C-D) did not align with each other. Instead, they were found to align to a genomic region located 3 megabases (Mb) distant (eFigure 3 E-F and G-H). This pattern of alignment suggests the presence of a complex genomic variant, denoted as DUP-NML-INV/DUP resulting in quadruplication of exon 4 and duplication of exon 3.(Figure 3). To validate this finding, we amplified the breakpoints (JC1 and JC2 in Figure 3), which are unique to this individual. Through this, we confirmed that this set of breakpoints only exists in this individual, hence they are not present in control samples (eFigure 4).

**Figure 3.**
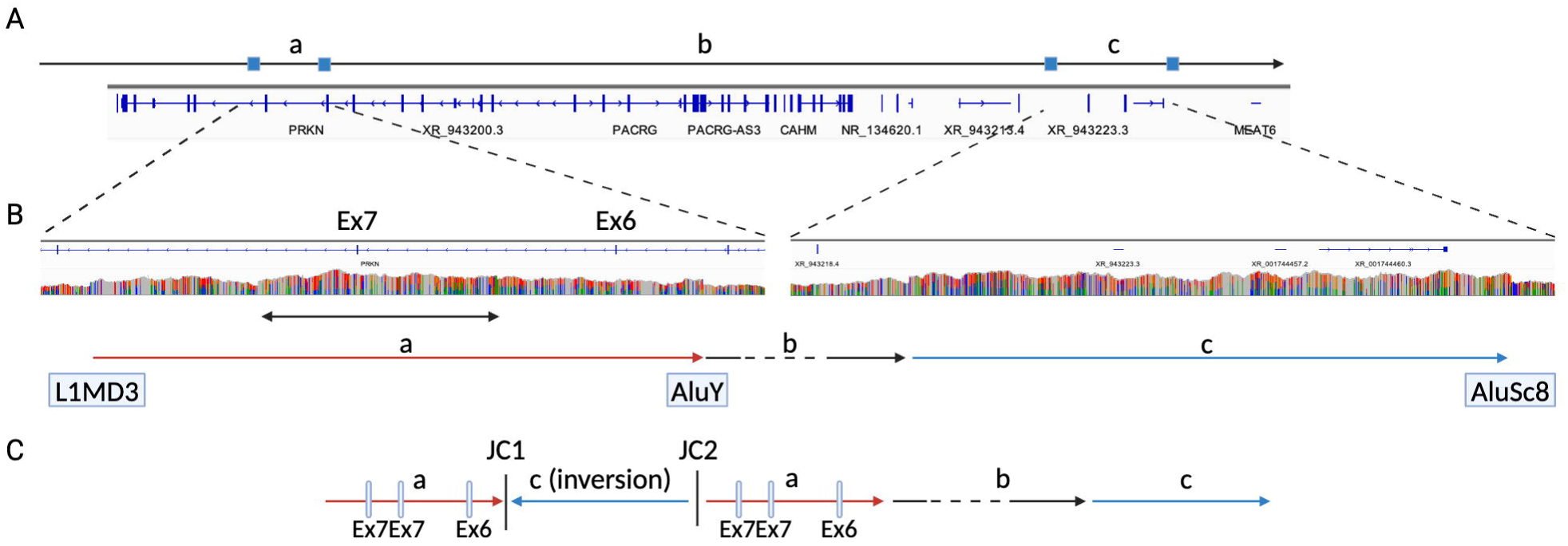
Description for the complex variant including exon 6 and 7 of PRKN. (A) Depicts the region of interest within the PRKN gene. Subregions (a-c) are indicated and correspond to the arrows marked a-c in panels B and C, highlighting specific areas of genetic variation. (B) Presents IGV screenshots illustrating the breakpoints associated with the complex variant. Notably, a multiplication of exon 7 is evident (bidirectional arrow), overlapped by an increased number of reads at arrow (a) including exon 6 and 7. Additionally, a multiplication within region (c) is also observed, showcasing the variant’s complexity. (C) Provides a schematic representation of the complex variant structure labeled as DUP-NML-DUP/INV. IGV; Integrative Genomics Viewer, INV; inversion, DUP;duplication, NML; normal

Considering inversion from PRKN-10, inversion was found in 4.35% (1/23) of the heterozygous *PRKN* carriers in the Japanese population. All the variants identified by long-read sequencing in this study are summarized in eTable 7.

### Clinical symptoms of long-read diagnosed *PRKN*-PD

Clinical symptoms of the six “new” two-variants *PRKN* cases are summarized in eTable 8. All patients except PRKN-26 presented typical presentations of *PRKN*-PD, showing AAO younger than 40 (AAO; 29.7 ± 14.84), normal heart-to-mediastinum (H/M) ratio in ^123^I-metaiodobenzylguanidine (MIBG) myocardial scintigraphy (75% (3/4)), less common autonomic symptoms (constipation 33% (2/6), urinary disturbance 0% (0/6), orthostatic hypotension 33% (2/6)), good response to levodopa (100% (6/6)), and less frequent olfactory dysfunction (0% (0/6)). PRKN-25, who harbored duplications of exon 5 and exon 5-6 had an AAO 40s and a family history of progressive supranuclear palsy (father). This patient had a decreased H/M ratio on MIBG myocardial scintigraphy, various autonomic symptoms, and levodopa equivalent dose of 1300mg 10 years after onset, which is atypical for *PRKN*-PD.

We then compared the clinical features between *PRKN* two-variant carriers and *PRKN* one-variant carriers. Five features showed different trends between one- and two-variant carriers (AAO, disease duration, gait disturbance, dystonia showing response to levodopa, and dystonia at onset). For example, the age of onset was younger in two-variant carriers (one-variant vs two-variants; 38.0 ± 7.16 vs 28.6 ± 11.32, p value = 0.0064) (eTable 9-10, eFigure 5).

### Breakpoints of pathogenic SVs

All the breakpoints and the locations of pathogenic SVs of *PRKN* identified in this study are summarized in Figure 4 and eTable 11. *PRKN* is located in one of the common fragile sites (CFS) in the genome, namely FRA6E, which makes the *PRKN* gene prone to have SVs. CFS are vulnerable to replication stress and often cause DNA breakage in this region, characterized by late replication, paucity of replication origins, and the ability to form DNA secondary structures.[30] In addition, Figure 4 also presents the core of FRA6E, as defined by Denison et al. using BAC clones RPCI-1 119H20 and RPCI-1 179P19. Since the precise location of RPCI-1 179P19 was unavailable, we used D6S1599 to represent the core visually.[31] All identified pathogenic SVs, except for two variants, had at least one breakpoint located in the core of FRA6E. (95%, 38/40) The two other variants were exon 1 deletion and exon 2 deletion of *PRKN*.

**Figure 4.**
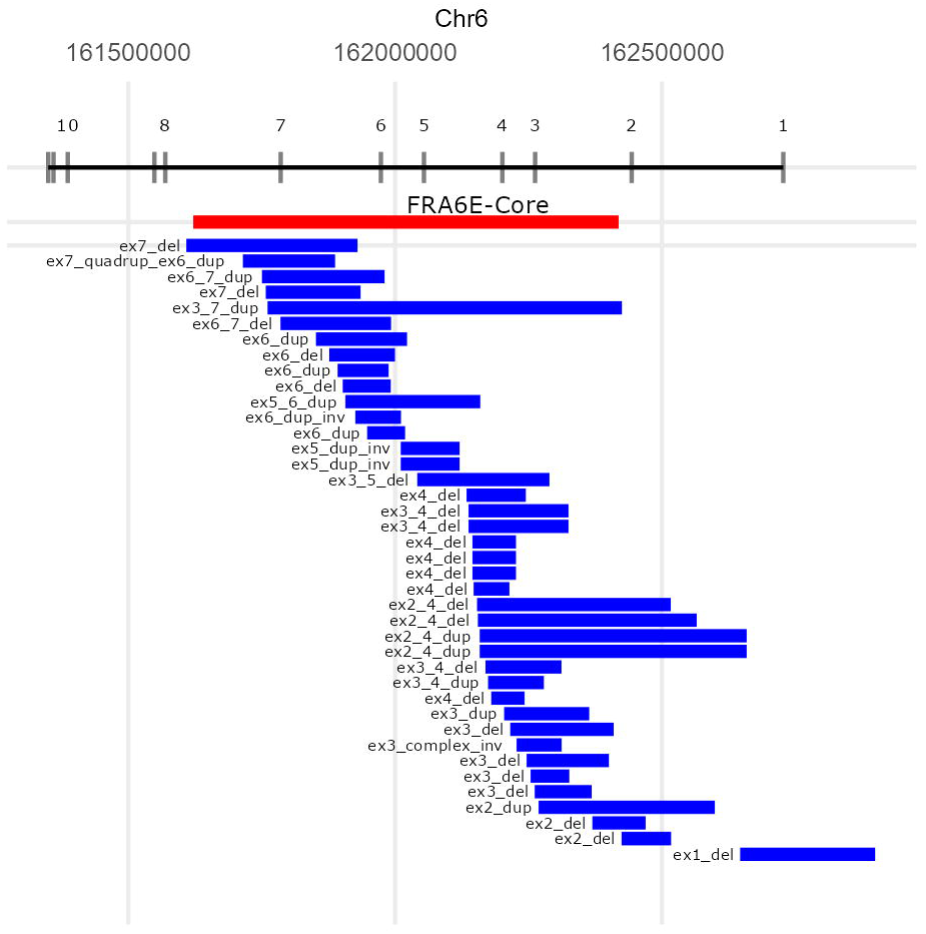
Genomic location of all the structural variants of *PRKN* identified in this study. Upper panel shows the location of exons of *PRKN*. Middle panel shows the location of the central core of FRA6E. The lower panel shows the location of all the SVs overlapped with exons of *PRKN* identified in this study. ex; exon, del; deletion, dup; duplication, dup-inv; duplication-inversion

## Discussion

In this study, we explored the performance of long-read sequencing for the identification of SNV and complex SV in the *PRKN*/*PINK1* genes. We included 12 known two-variants *PRKN*-PD as “positive controls” and all 24 variants were successfully identified. Next, we wanted to identify complex and previously undetected secondary variants in *PRKN*/*PINK1* heterozygous carrier patients, potentially demonstrating that complex SVs and overlapping SVs of *PRKN* are likely missed by traditional sequencing methods and MLPA. In our cohort, we could identify a second variant in 26% (n=6) of the *PRKN* heterozygous carriers and in 0% of the *PINK1* heterozygous carriers. This study shows the utilization of long-read sequencing in the diagnosis of EOPD and long-read sequencing should be considered as a next step after short-read sequencing and MLPA for unresolved EOPD cases,

Approximately 5-10% of PD patients can be classified as monogenic PD, which means a single gene is mainly responsible for their disease development. In our study using short-read targeted resequencing for PD related genes in the EOPD population, surprisingly, 60% of these patients remained undiagnosed. However, our research indicates that using long-read sequencing could be more effective. In fact, more than 20% of patients with a single variant in the *PRKN* gene were successfully diagnosed with this method in this study. Therefore, for those patients who remained undiagnosed after short-read sequencing, long-read sequencing might provide a diagnosis. Given that *PRKN* along with *GBA1* and *LRRK2*, are targets for gene therapy in PD, applying long-read sequencing to the EOPD population may offer a broader range of candidates for the upcoming gene-therapy era[32]. We anticipate that the utilization of long-read sequencing will become more widespread in the diagnosis of familial PD, particularly in unresolved high suspect monogenic/early onset cases.

An inversion was detected in 4.35 % (1/23) of the heterozygous *PRKN* carriers in the Japanese population. Considering the case of the massive inversion we recently reported, it is suggested that inversions in PRKN are not an extremely rare type of SV.[15] We have also identified another complex SV, DUP-NML-INV/DUP which included *PRKN* exons (Figure 3). MLPA called a duplication or triplication of *PRKN* exon 7 but did not call a CNVs in exon 6 (eFigure 2). For this variant, two of the junctions overlapped with an *Alu* transposable element in the reference genome, which is in line with the previous reports that described DUP–NML–INV/DUP was mediated by *Alu-Alu* rearrangements (Figure 3).[33–35] This case underscores the complexity of SVs in *PRKN*.

These cases highlight the utility of long-read sequencing and we believe that long-read sequencing should be considered as a next step after short-read sequencing and MLPA for unresolved EOPD cases, especially with a heterozygous *PRKN* variant. Moreover, although it may not be frequent, we may need to consider that there should be EOPD cases with *PRKN*-PD phenotype harboring homozygous variants of complex SVs of *PRKN* when short read sequencing could identify any pathogenic variant.

In addition to this study, three cases of a pathogenic inversion involving *PRKN* have been reported.[15,36,37] One case is from Israel, describing EOPD patients with homozygous 77 Kb inversion including exon 5 from consanguineous family. Second, our case from Japan showed monozygotic twins with compound heterozygous *PRKN* variants of exon three deletion and 7 Mb inversion including exon 1 to 11. The last case was from Poland, describing inversion including exon 2 to exon 5, which was a part of duplication. Given the observation of *PRKN* inversion across various populations, including Jewish, European, and Asian, it is reasonable to infer that this genetic variation can be identified in a wide range of ethnic backgrounds. We may need to sequence a larger number of samples from diverse populations to know the frequency of inversions of *PRKN*.

Long-read sequencing also helped identify the variants of *PRKN* when the SVs of each allele overlap or when the SVs are contiguous. Three out of six long-read diagnosed *PRKN*-PD cases harbored overlapping or contiguous SVs. We have previously reported that overlapping of a deletion and duplication in the same allelic exon could be normal in qPCR and differentiated them using parental DNA.[38] It is natural to consider that overlap of deletion and duplication can be missed by MLPA. In this study, we identified exon 2 deletion, exon 3-4 duplication in one allele and exon 4 deletion in another allele (PRKN-11) by long-read sequencing (eFigure 2). Using qPCR and MLPA, it was observed that only exon 2 was deleted (duplication of exon 3 was not consistent between qPCR and MLPA). Parental DNA was needed to phase the variants in conventional methods, however, using long-read sequencing, it is able to differentiate and phase the overlapped variants only by proband’s DNA. Similar phenomenon was observed in PRKN-18 from *PRKN* two variants group, who had known to have exon 2 duplication and exon 5 deletion but long-read sequencing revealed the true variant to be exon 2-4 duplication and exon 3-5 deletion. Long-read sequencing was also useful in distinguishing between two deletions in continuous exons, which appeared to be one deletion including two exons by MLPA. (PRKN-33). These findings underlied the utility of long-read sequencing on accurate diagnosis of *PRKN*-PD when SVs are overlapped or continuous.

When we compared the clinical phenotype of PRKN two variants group (n=18) and PRKN single variant group (n=18), five features had a significant correlation with the number of the PRKN variants (eTable 10, eFigure 5). AAO was younger in the two-variants group, suggesting the true PRKN-PD patients are likely to have younger AAO. On the other hand, disease duration was shorter in the one variant group. It may cause inaccurate diagnosis of PD in those patients.In this study we could not find any SVs in *PINK1*. Reported SVs of *PINK1* include deletion of single exon, multiple exons or whole gene in multiple populations, and exonic duplications.[14] We hypothesized that there may be complex SVs as a hidden variant but none was identified. The reason complex SVs were not identified could be due to them being truly absent, or due to the small number of samples (n = 12). We plan to perform long-read sequencing to larger samples to elucidate the presence of complex SVs in *PINK1* in the future.

A key question that can be addressed from the findings of this study is: when do we need to consider long-read sequencing for EOPD? An important consideration is that MLPA can be performed for $20 per sample, whereas long-read sequencing with ONT costs approximately $1000 per sample. It is reasonable, therefore, to perform MLPA to screen for SVs first.[39] In our previous study, we combined MLPA with targeted resequencing/Sanger sequencing to analyze EOPD patients (n = 918) with an AAO younger than 50 years in a Japanese cohort. We identified that 6.4% of the patients harbored two variants in the PRKN gene, while 3.9% presented with a single variant.[9] A study from the United Kingdom reported 2.3% of two variant carriers of PRKN and 3.8% of single variant carriers from EOPD with AAO younger than 50 using direct sequencing and MLPA.[40] In addition, a recent paper showed *PRKN*-PD is more common (18 per 100,000 individuals) than it has been thought (35,000 - 70,000 worldwide), which suggests the number of PD patients with *PRKN* variant should be larger.[41,42] Thus, it is assumed that there is a certain number of EOPD patients with heterozygous *PRKN* variants after checking pathogenic SNVs and CNVs by conventional methods, which is considered to be a good application for long-read sequencing, as we did in this study.

To our knowledge, this is the first large-scale study to apply long-read sequencing to *PRKN* SVs describing the break points of pathogenic SVs more accurately. Importantly, almost all *PRKN* SVs we identified in this study were located in the central core of FRA6E (Figure 4).[31] When we compared the location of SVs identified in this study to the SVs recurrently observed in the study from Mitsui et al, four SVs were common. These four SVs were found from Japanese or Asian populations in their study but not from European populations. These facts support the necessity of long-read sequencing for the identification of complex SVs, as it seems to be difficult to identify a region in which complex SVs frequently occur and screen them using cheaper techniques like Sanger sequencing. Moreover, as we confirmed that pathogenic SVs of *PRKN* were concentrated in the FRA6E core. We speculate that looking closer to the SVs in common fragile sites may lead us to identify more disease or phenotype related SVs in neurodegenerative disease, especially in familial cases.

This study has some limitations. First, the current sample size is large for *PRKN*-PD but still limited in order to know the true frequency of complex SVs like inversion. More long-read sequencing data is needed including large numbers of controls to know the frequency of inversions across populations. Second, we were not able to confirm the changes of RNA transcripts in samples with complex SVs in *PRKN*. Attempts using RT-PCR and RNAseq from mRNA extracted from peripheral blood were not successful due to the low expression of PRKN and unfortunately no other patient material is available. Third, we only had access to samples from Japanese ancestry. A more diverse population is needed to know the true significance of complex SVs in EOPD. We are now in the process of applying long-read sequencing to different ancestral populations.

## Conclusions

In summary, this study was the first study to use long-read sequencing on a large group of EOPD patients to identify hidden and complex SVs. This study demonstrated the complexity of SVs in the *PRKN* gene which is even more complex than previously thought. Additionally, the study highlighted the effectiveness of long-read sequencing in researching the genetics of EOPD. It is expected that the application of long-read sequencing will increase, leading to more accurate and faster diagnoses which is important for *PRKN*-PD given the potential need for genetic counseling, different progression vs idiopathic PD and eligibility for clinical trials.

## Supporting information

eFigure1

eFigure2

eFigure3

eFigure4

eFigure5

eTable1

eTable2

eTable3

eTable4

eTable5

eTable6

eTable7

eTable8

eTable9

eTable10

eTable11

## Data Availability

All the data publicly available is included in the manuscript. The authors declare that the genomics data in this study is not available to share due to the ethics.

## List of abbreviations

PD: Parkinson’s disease
GWAS: genome-wide association studies
EOPD: early onset Parkinson’s disease
AAO: age at onset
SNV: single nucleotide variant
MLPA: multiplex ligation-dependent probe amplification
SV: structural variant
CNV: copy number variant
qPCR: quantitative PCR
ONT: Oxford Nanopore Technologies
IGV: integrative genome viewer
H/M: heart-to-mediastinum
MIBG: 123I-metaiodobenzylguanidine
CFS: common fragile sites

## Declarations

### Ethics approval and consent to participate

The study was approved by the ethics committee of Juntendo University, Tokyo, Japan, and all participants provided written informed consent to participate in the research described in this study (M08-0477-M09).

### Consent for publication

All participants provided written informed consent to publish the research described in this study (M08-0477-M09).

### Competing interests

The authors declare that they have no competing interest.

### Funding

This research was supported in part by the Intramural Research Program of the NIH, National Institute on Aging (NIA), National Institutes of Health, the Japan Society for the Promotion of Science (JSPS) KAKENHI [Grant Numbers: 24K02372 and 23K06958 for MF, 22K07542 for HY, 21K07283 for YL, 20K07893 for KN, 21H04820 and 24H00068 for NH], the Japan Science and Technology Agency (JST) Moonshot R&D Program [Grant Number: JPMJMS2024-5 for NH], AMED under Grant Number 23bm1423015h0001 for MF and NH, and 24ek0109677h0002 for NH, Subsidies for Current Expenditures to Private Institutions of Higher Education from the Promotion and Mutual Aid Corporation for Private Schools of Japan, through a subaward from Juntendo University (for MF and NH), and the Research Institute for Diseases of Old Age, Juntendo University Graduate School of Medicine (for MF, YH, and NH).

### Author’s contributions

Concept and design: KD, MF, CB, NH

Acquisition, analysis, or interpretation of data: KD, HY, LM, BB, RG, KP, MI, MF, YI, KN, SM, MH, KT, KJB, MF, CB

Drafting of the manuscript: KD, KJB, MF, CB

Critical review: All

Supersision: CB, NH

## Acknowledgements

We thank all the participants who contributed to this study. We thank the Biowulf team, as this study used the high-performance computational capabilities of the Biowulf Linux cluster at the National Institutes of Health (http://hpc.nih.gov). This work was in part supported by the Intractable Disease Research Center of Juntendo University Graduate School of Medicine.

Figures 1 and 2 were generated on www.biorender.com.

KD reports receiving grants from the JSPS Research Fellowship for Japanese Biomedical and Behavioral Researchers at NIH.

HY reports receiving grants from the JSPS (22K07542).

LM reports no disclosures.

BB reports no disclosures.

RG reports no disclosures.

KP reports no disclosures.

MI reports no disclosures.

YL reports receiving grants from the JSPS (21K07283).

KN reports receiving grants from the JSPS (20K07893).

SM reports no disclosures.

MH has received funding for speaker honoraria from Sumitomo, Ono, Otsuka, Novartis, Kyowa-Kirin, Eisai, and Takeda, and received research funding that is not related to this study from Takeda.

KT reports no disclosures.

KB reports no disclosures.

MF reports grants from Japan Agency for Medical Research and Development GAPFREE (21ak0101125h0002); Subsidies for Current Expenditures to Private Institutions of Higher Education from the Promotion and Mutual Aid Corporation for Private Schools of Japan.

CB reports no disclosures.

NH reports receiving the following grants and fees unrelated to this research during the conduct of the study: grants from the Japan Society for the Promotion of Science (JSPS), the Japan Agency for Medical Research and Development (AMED), the Japan Science and Technology Agency (JST) Moonshot R&D Program (JPMJMS2024-5), the Japan Science and Technology Agency (JST), a Health Labour Sciences Research Grant, IPMDS, and MJFF; personal fees and speakers’ honoraria from Sumitomo Pharma, Takeda Pharmaceutical, Kyowa Kirin, AbbVie GK, Otsuka Pharmaceutical, Novartis Pharma, Ono Pharmaceutical, Eisai, Teijin Pharma, and Daiichi Sankyo Co. FP Pharma; personal fees for consultants and advisory boards from Sumitomo Pharma, Takeda Pharmaceutical, Kyowa Kirin, Ono Pharmaceutical, Teijin Pharma, and PARKINSON Laboratories Co.; and he owns shares in the PARKINSON Laboratories Co. Ltd (Equity stock (8%)).

Dr Blauwendraat and Dr Hattori had full access to all of the data in the study and take responsibility for the integrity of the data and the accuracy of the data analysis

During the preparation of this work the authors used ChatGPT4 in order to improve language. After using this tool/service, the authors reviewed and edited the content as needed and take full responsibility for the content of the publication.

## Additional files

**eFigure1**

eFigure1.pdf

Histogram of data outputs of long-read sequencing

**eFigure2**

eFigure2.pdf

Comparison of MLPA and Long-read sequencing in all *PRKN* variant carriers

**eFigure3**

eFigure3.pdf

Screenshot of IGV for DUP-NML-DUP/INV

The upper panel showed the region including exon six and seven of *PRKN*. The lower panel showed the region 3 Mb away from the upper panel. The reads mapped at two breakpoints (A-B and C-D) of the upper panel were split but not aligned to each other. The split reads were aligned in the breakpoints in the lower panel (E-F and G-H).

IGV; integrative genome viewer, DUP; duplication, NML; normal, INV; inversion

**eFigure4**

eFigure4.pdf

PCR for the junctions of DUP-NML-DUP/INV

**eFigure5**

eFigure5.pdf

Correlation between the number of *PRKN* variants number and clinical phenotype

**eTable 1**

eTable1.csv

Primers for Sanger sequencing

**eTable 2**

eTable2.csv

Results of DNA quality control checking for long-read sequencing

**eTable 3**

eTable3.csv

*PRKN* variant carriers by MLPA and short-read sequencing

**eTable 4**

eTable4.csv

*PINK1* variant carriers by MLPA and short-read sequencing

**eTable 5**

eTable5.csv

Long-read sequencing data outputs

**eTable 6**

eTable6.csv

Long-read sequencing adaptive sampling data outputs

**eTable 7**

eTable7.csv

All variants identified by long-read sequencing

**eTable 8**

eTable8.csv

Clinical phenotype of Samples with two *PRKN* variants only by long-read sequencing

**eTable 9**

eTable9.csv

Clinical phenotype of all *PRKN* carriers

**eTable 10**

eTable10.csv

Correlation of the number of *PRKN* variants and clinical phenotype

**eTable 11**

eTable11.csv

Breakpoints of *PRKN* structural variants

## Notes

### Competing Interest Statement

The authors have declared no competing interest.

### Author Declarations

The ethics committee of Juntendo University, gave ethical approval for this work (M08-0477-M09).

