## Supplementary figures and images for "The Utility of Long-Read Sequencing in Diagnosing Genetic Autosomal Recessive Parkinson’s Disease: a genetic screening study"

### eFigure1

**eFigure 1 Histogram of data outputs of long-read sequencing**

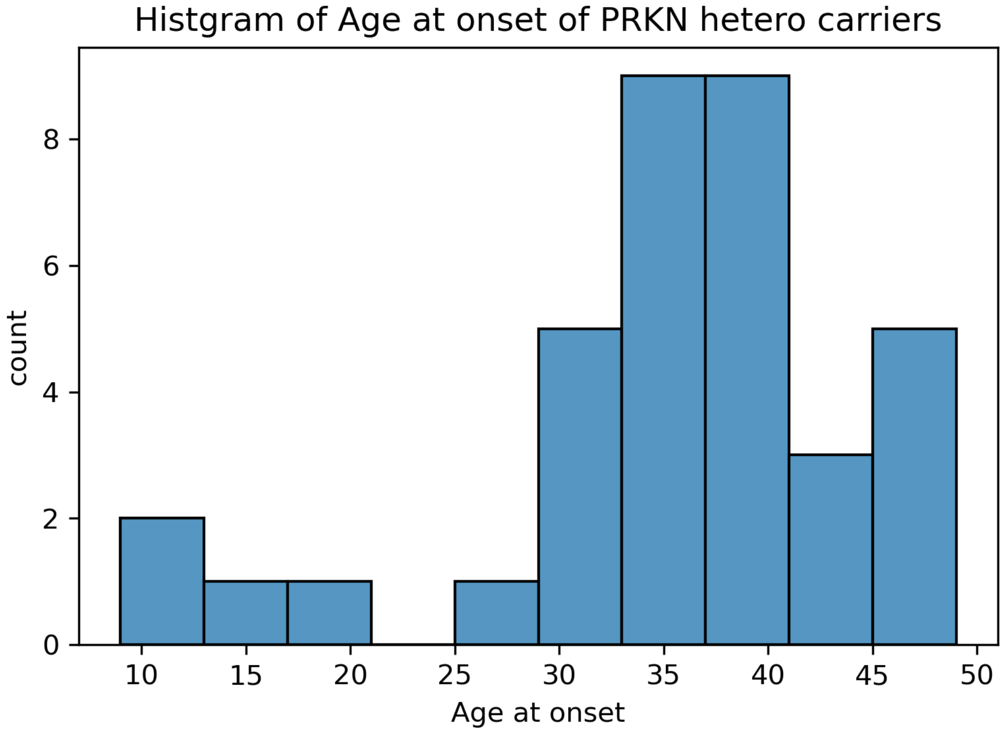


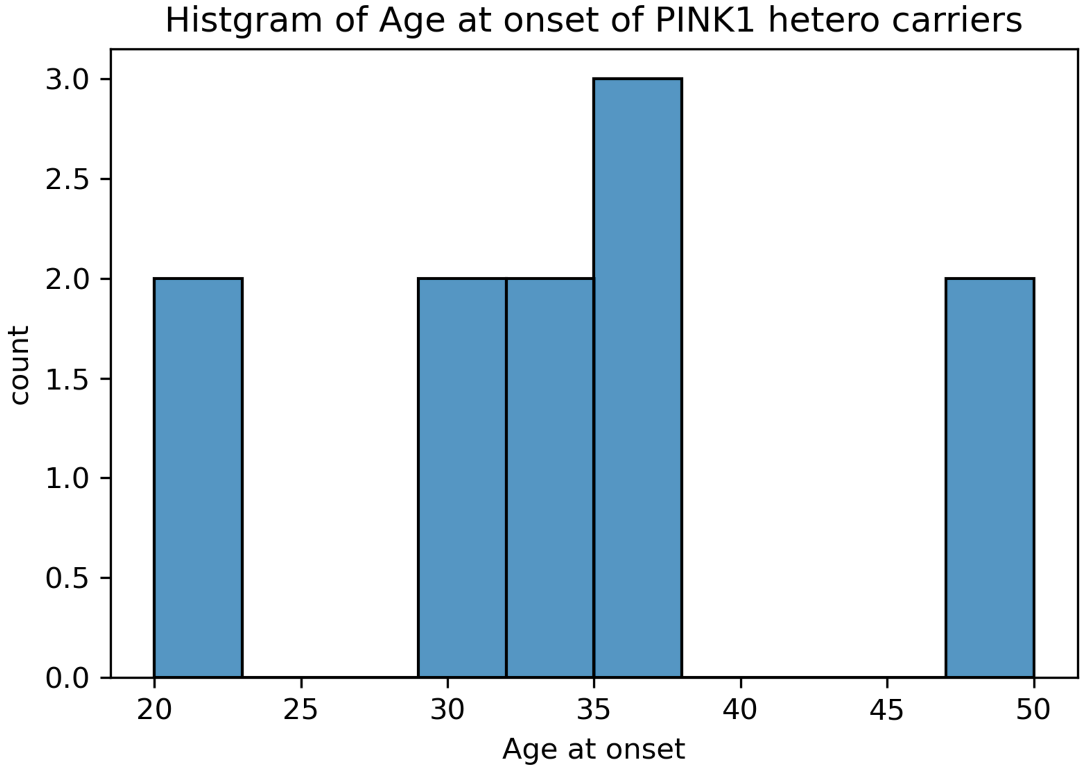
