## Supplementary material for "The Utility of Long-Read Sequencing in Diagnosing Genetic Autosomal Recessive Parkinson’s Disease: a genetic screening study": eFigure2

### eFigure 2 Comparison of MLPA and Long-read sequencing in all *PRKN* variant carriers

The header of each figure describes the sample name and the structural variants if they have.

PRKN-1: Exon 4 deletion and Exon 6 deletion

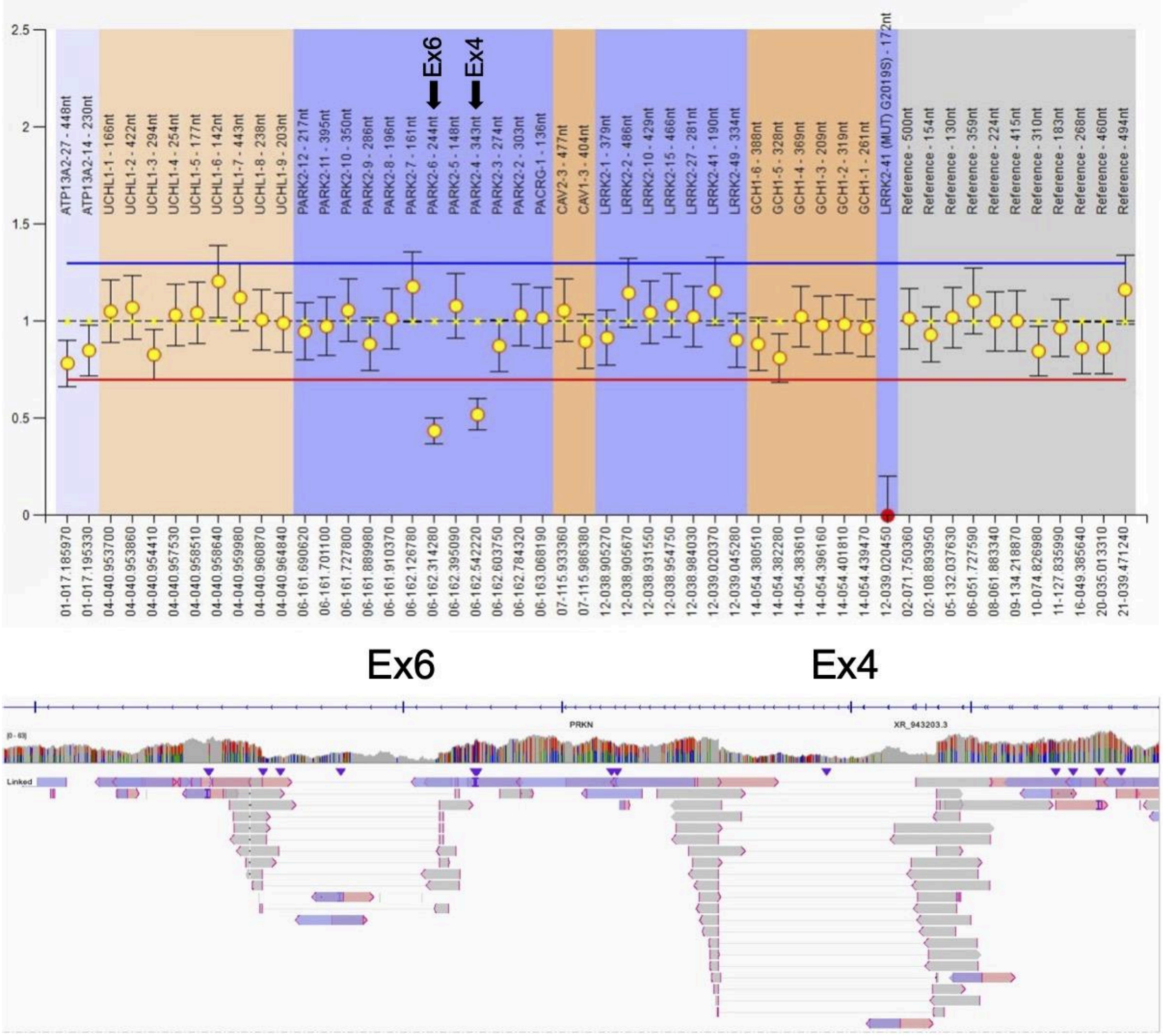

PRKN-2: Exon 2-4 deletion

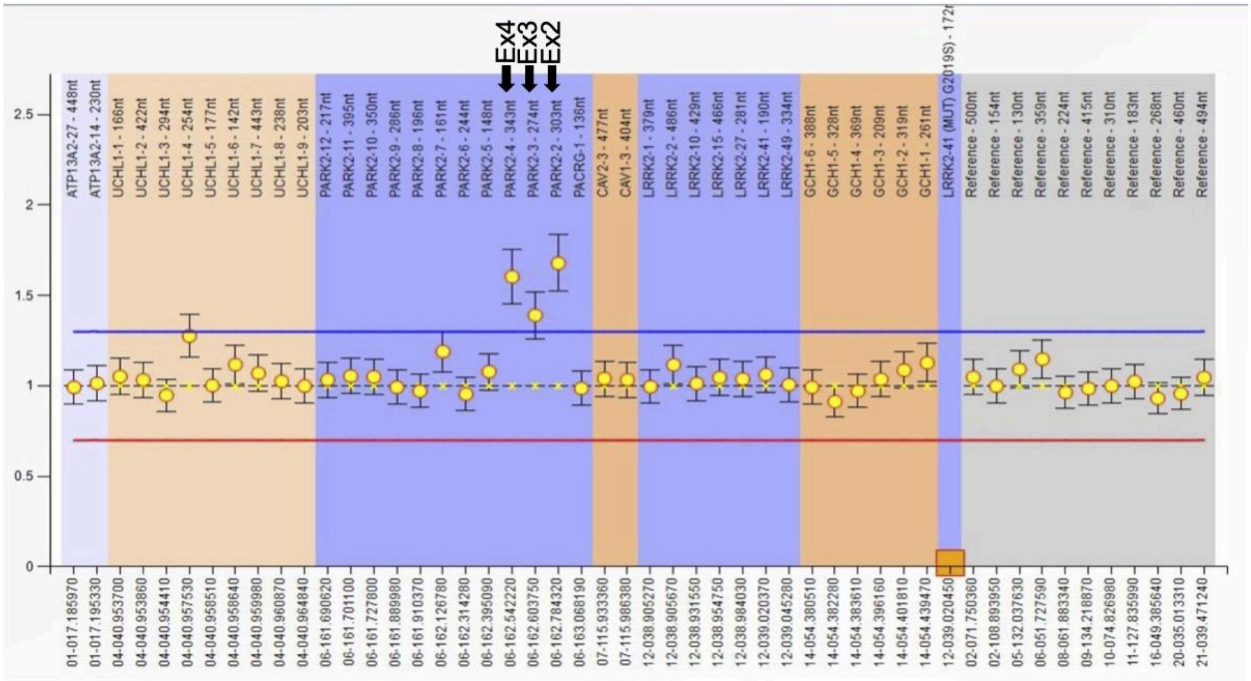


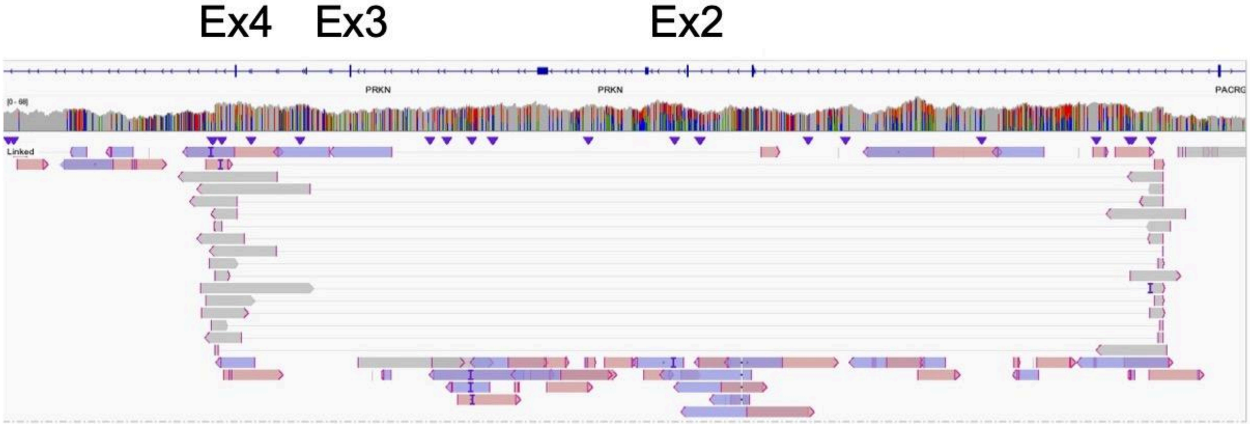

PRKN-3: Exon 6-7 deletion

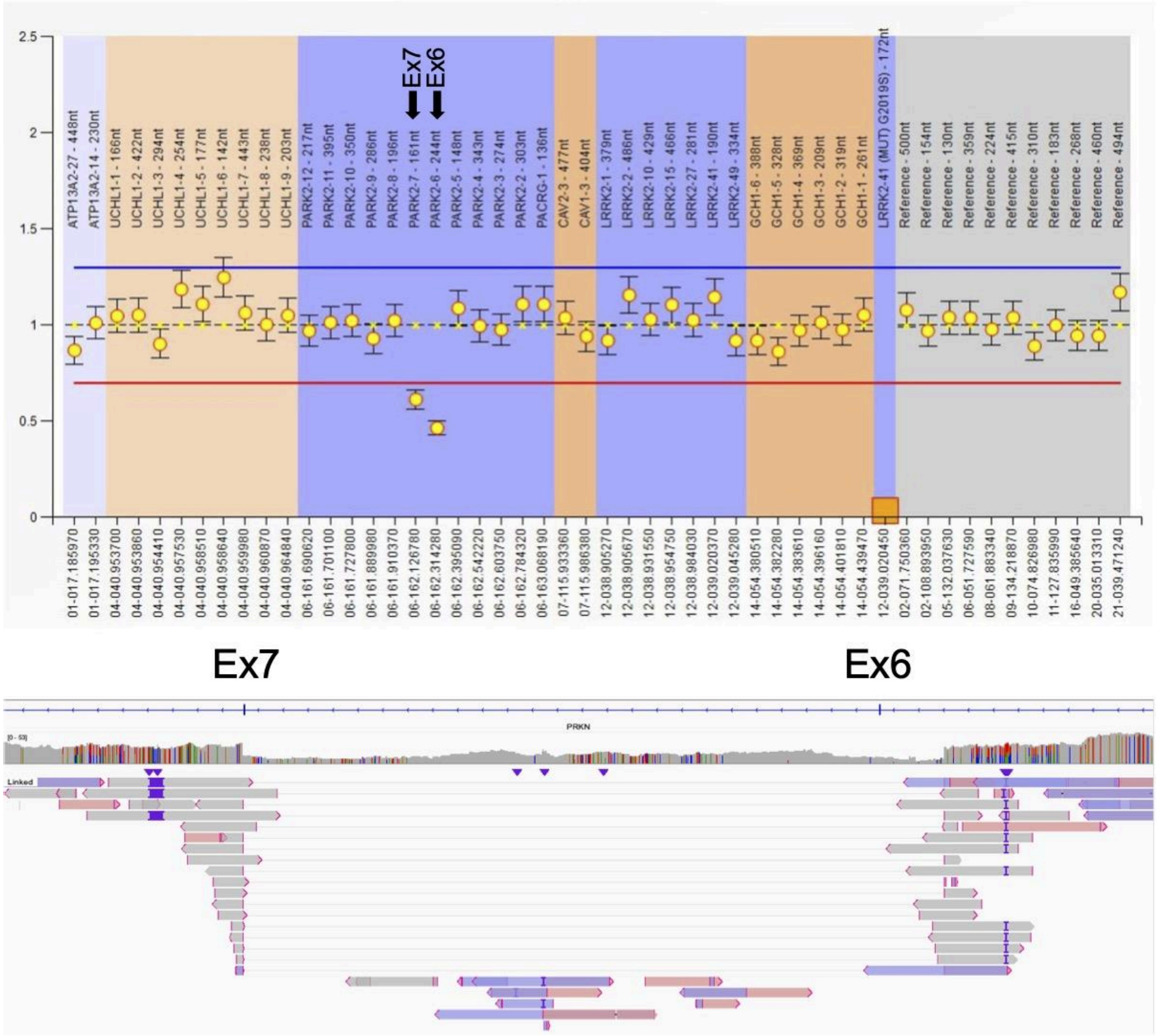

PRKN-4: Exon 4 deletion

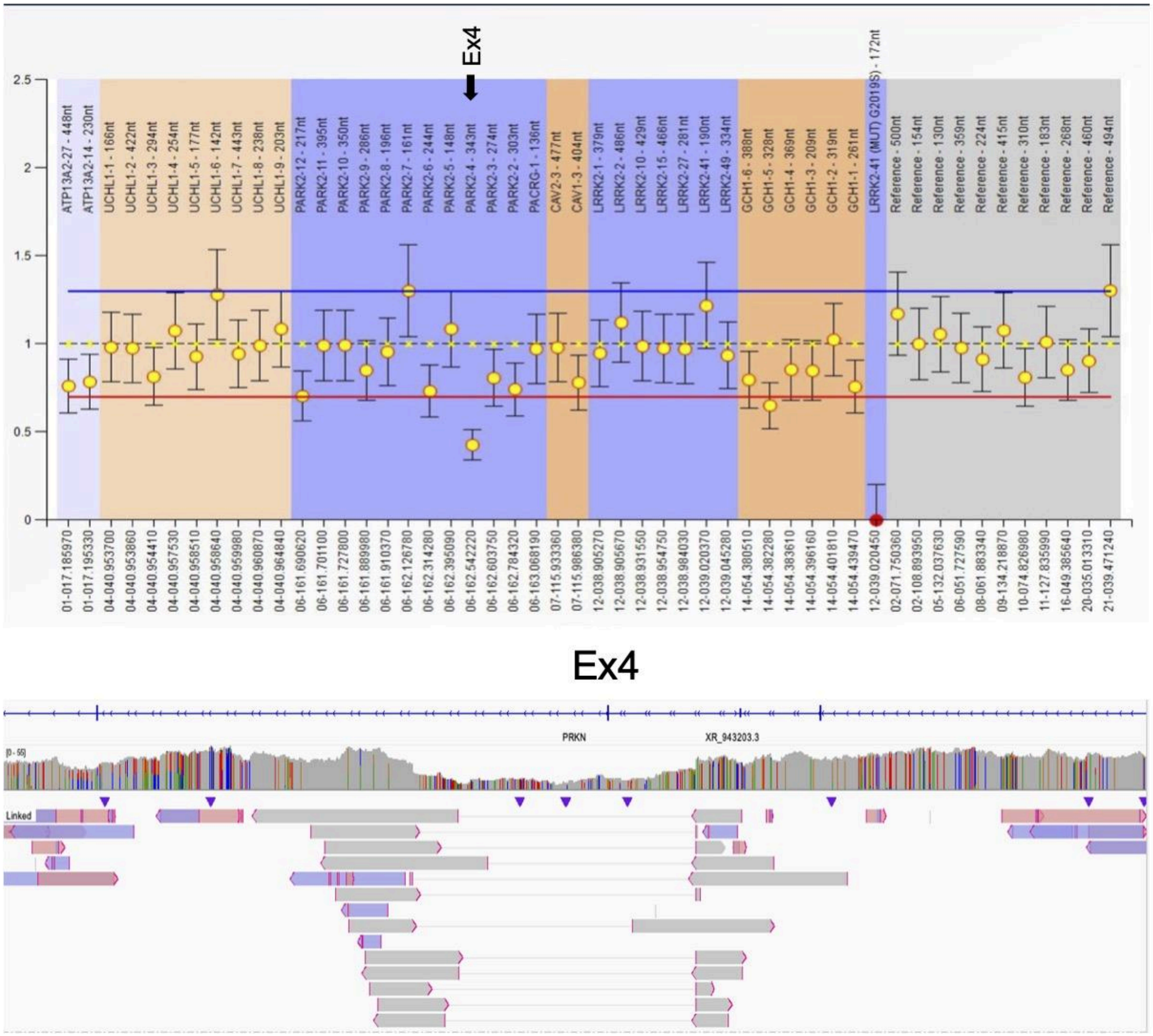

PRKN-5: Exon 2-4 duplication

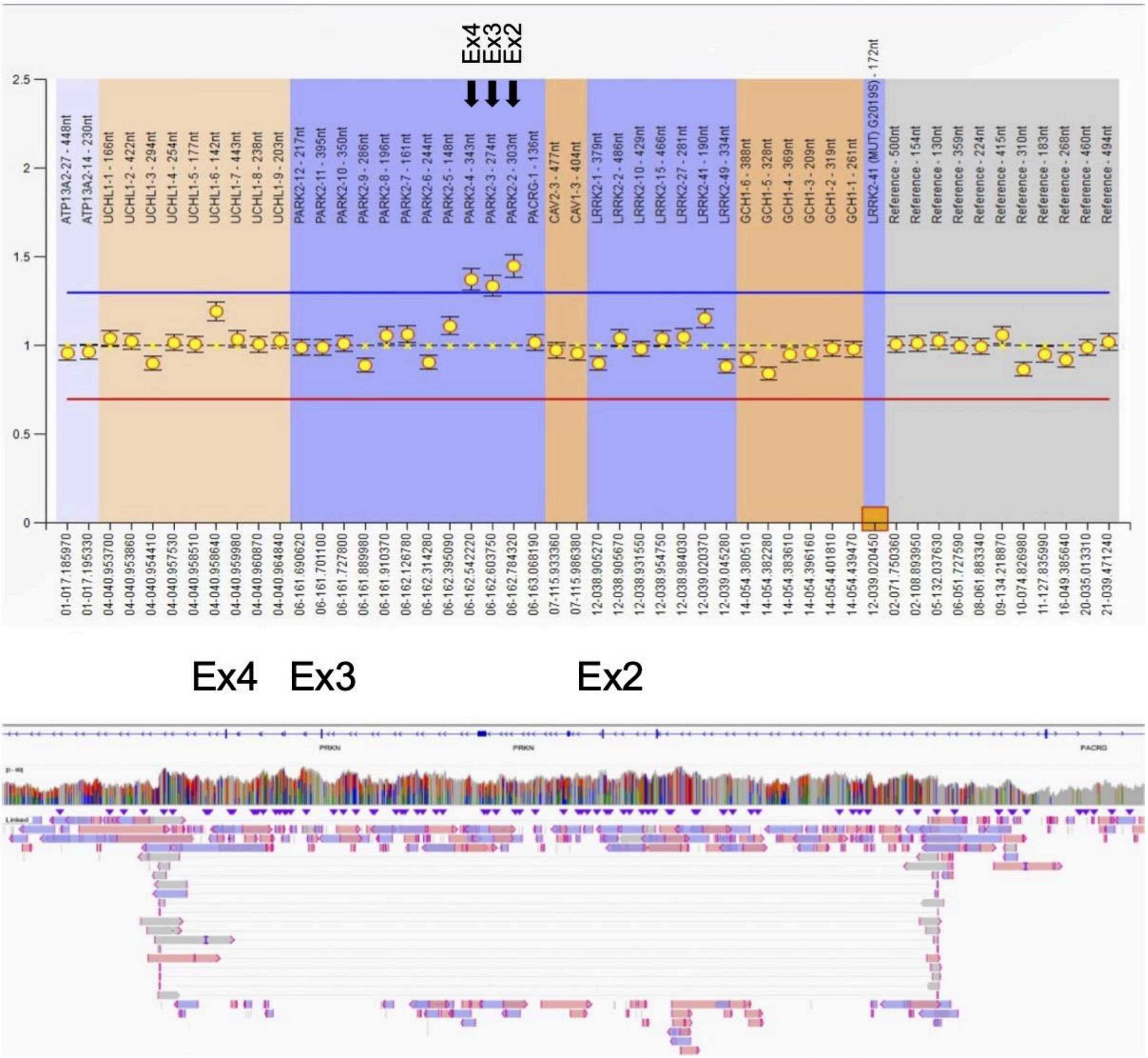

PRKN-6: Exon 4 deletion and Exon 3 duplication

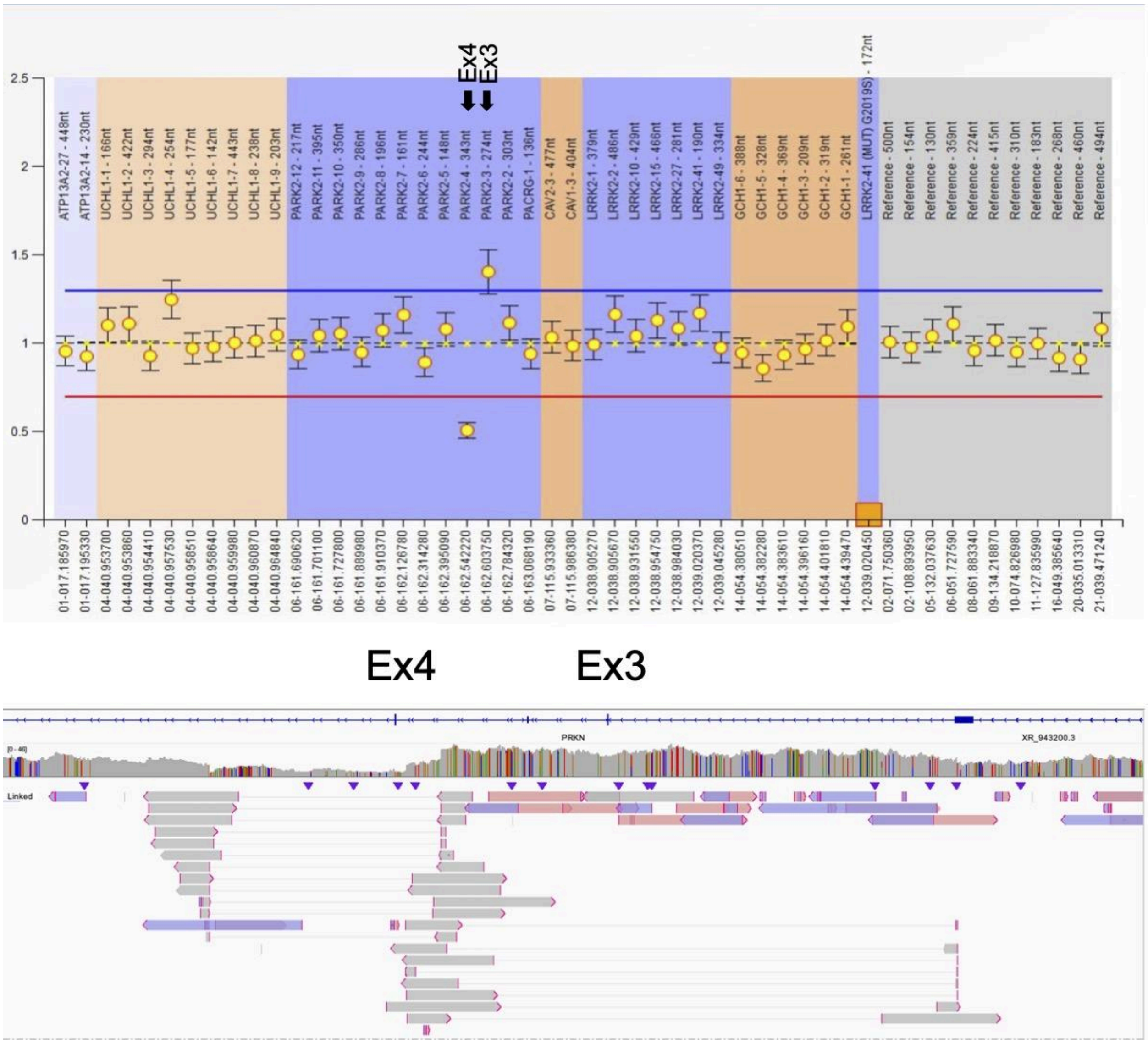

PRKN-7: Exon 4 deletion

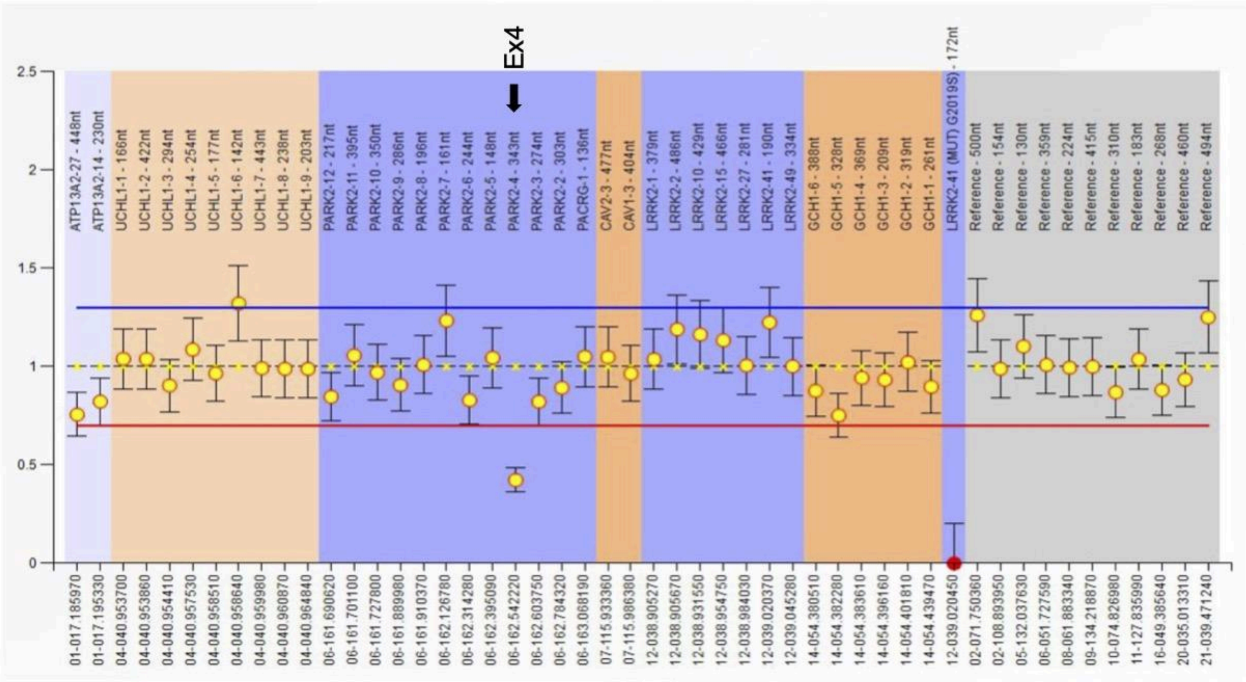


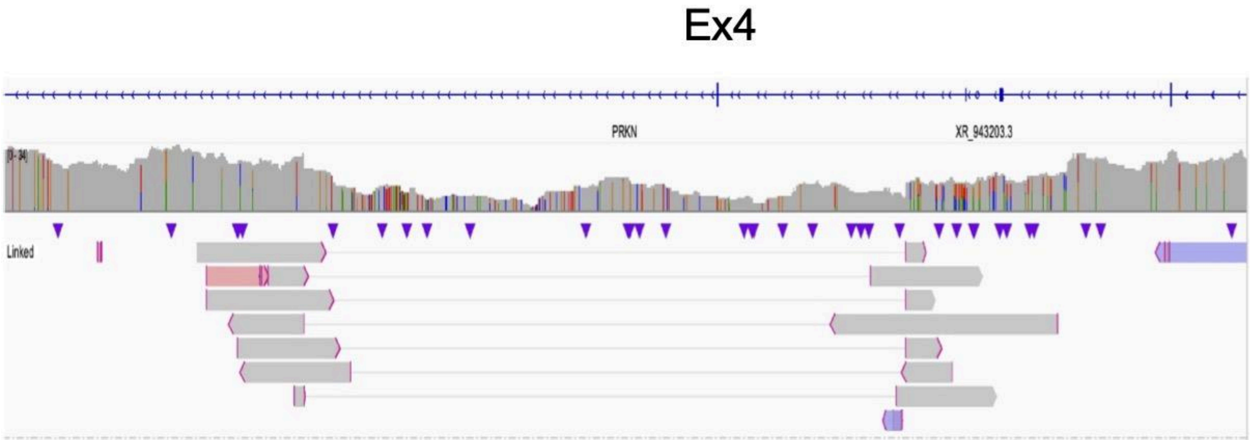

PRKN-8:

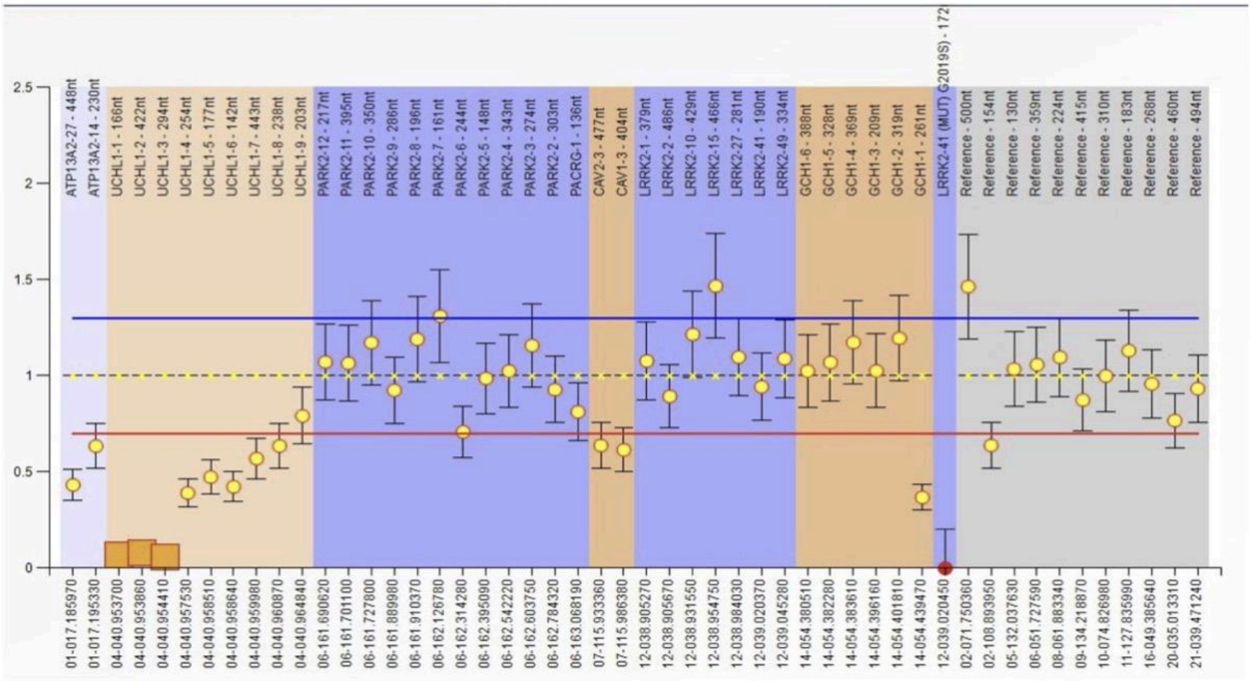

2.5
2
1.5
1
0.5
0
ATP13A2-27 - 448nt
ATP13A2-14 - 230nt
UCHL1-1 - 166nt
UCHL1-2 - 422nt
UCHL1-3 - 294nt
UCHL1-4 - 254nt
UCHL1-5 - 177nt
UCHL1-6 - 142nt
UCHL1-7 - 443nt
UCHL1-8 - 238nt
UCHL1-9 - 203nt
PARK2-12 - 217nt
PARK2-11 - 395nt
PARK2-10 - 350nt
PARK2-9 - 286nt
PARK2-8 - 196nt
PARK2-7 - 161nt
PARK2-6 - 244nt
PARK2-5 - 148nt
PARK2-4 - 343nt
PARK2-3 - 274nt
PARK2-2 - 303nt
PACRG-1 - 136nt
CAV2-3 - 477nt
CAV1-3 - 404nt
LRRK2-1 - 379nt
LRRK2-2 - 486nt
LRRK2-10 - 429nt
LRRK2-15 - 466nt
LRRK2-27 - 281nt
LRRK2-41 - 190nt
LRRK2-49 - 334nt
GCH1-6 - 388nt
GCH1-5 - 328nt
GCH1-4 - 369nt
GCH1-3 - 209nt
GCH1-2 - 319nt
GCH1-1 - 261nt
LRRK2-41 (MUT) G2019S) - 172
Reference - 500nt
Reference - 154nt
Reference - 130nt
Reference - 359nt
Reference - 224nt
Reference - 415nt
Reference - 310nt
Reference - 183nt
Reference - 268nt
Reference - 460nt
Reference - 494nt
01-017.185970
01-017.195330
04-040.953700
04-040.953860
04-040.954410
04-040.957530
04-040.958510
04-040.958640
04-040.959980
04-040.960870
04-040.964840
06-161.690620
06-161.701100
06-161.727800
06-161.889980
06-161.910370
06-162.126780
06-162.314280
06-162.395090
06-162.542220
06-162.603750
06-162.784320
06-163.068190
07-115.933360
07-115.986380
12-038.905270
12-038.905670
12-038.931550
12-038.954750
12-038.984030
12-039.020370
12-039.045280
14-054.380510
14-054.382280
14-054.383610
14-054.396160
14-054.401810
14-054.439470
12-039.020450
02-071.750360
02-108.893950
05-132.037630
06-051.727590
08-061.883340
09-134.218870
10-074.826980
11-127.835990
16-049.385640
20-035.013310
21-039.471240

PRKN-9: Exon 7 quadruplication and Exon 6 duplication (inside of DUP-NML-DUP/INV)

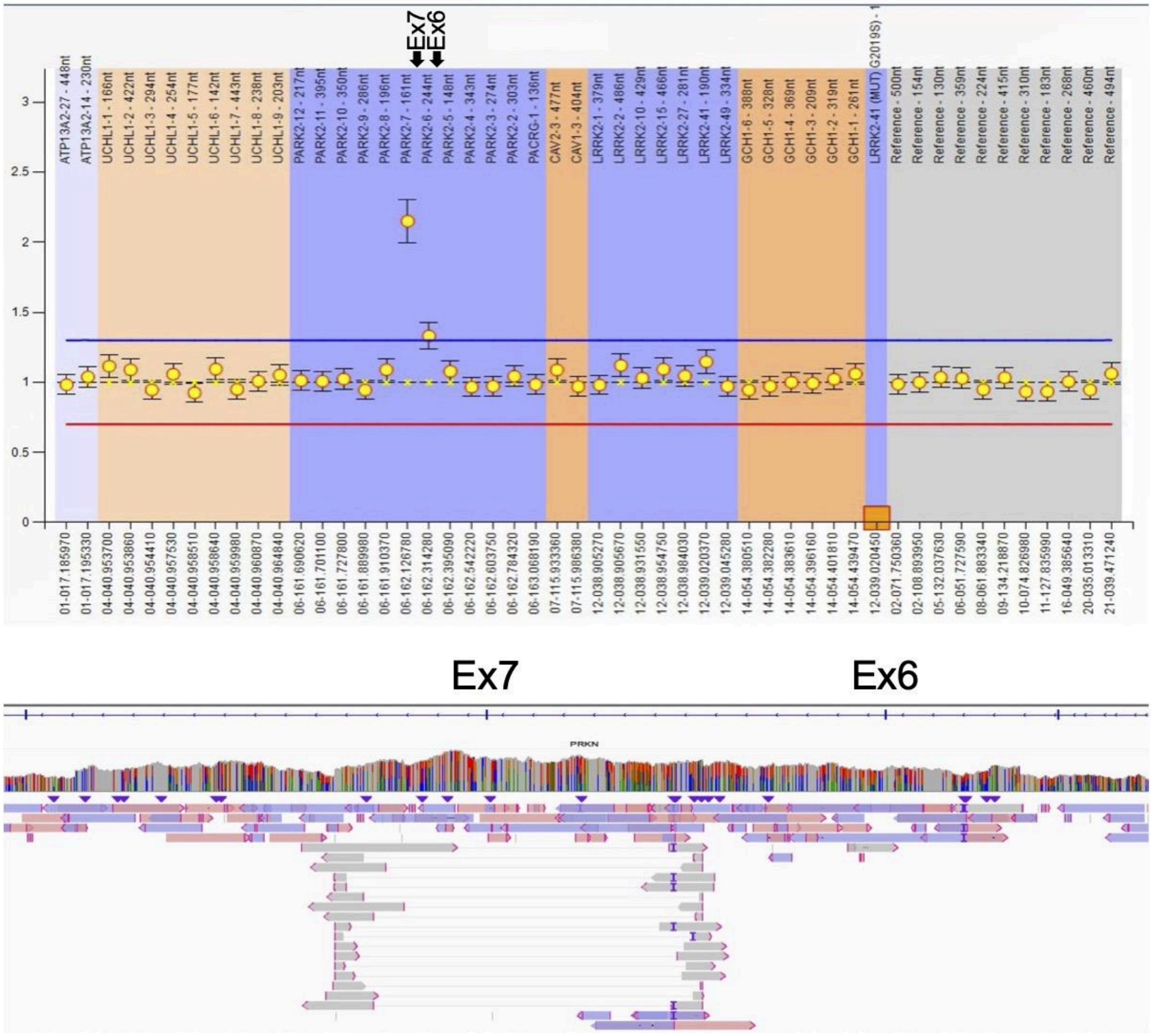

PRKN-10: Exon 3 deletion and Exon 3 complex inversion

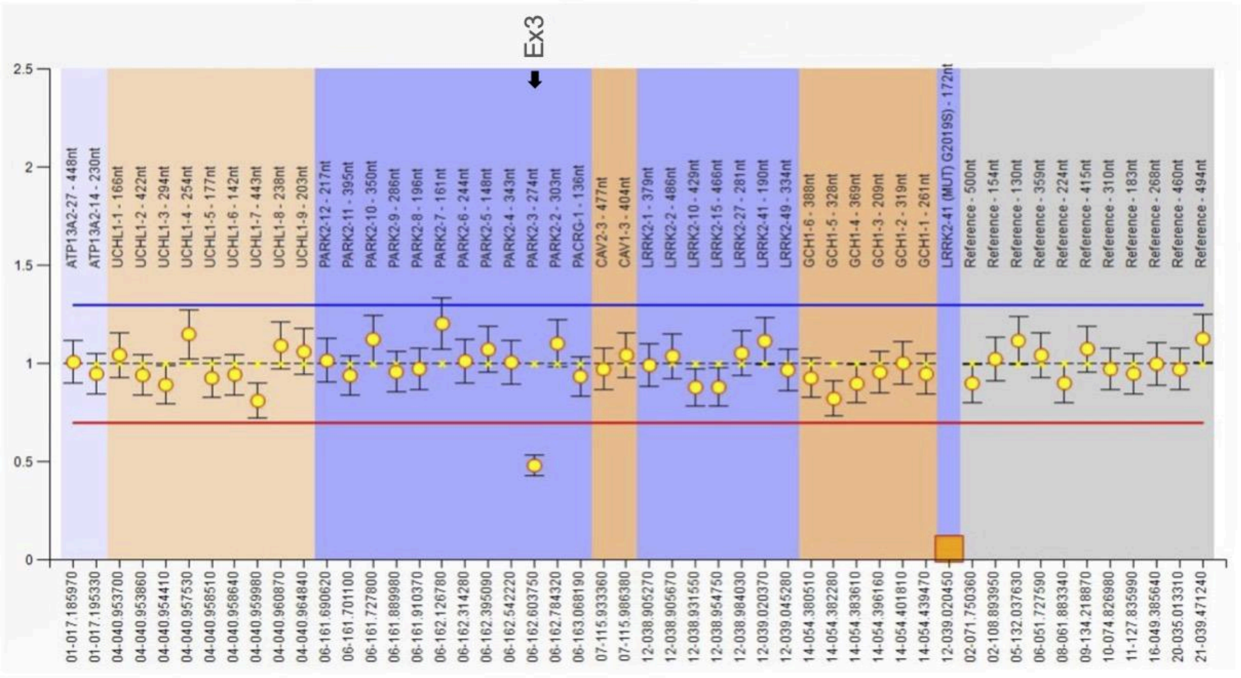


Ex3

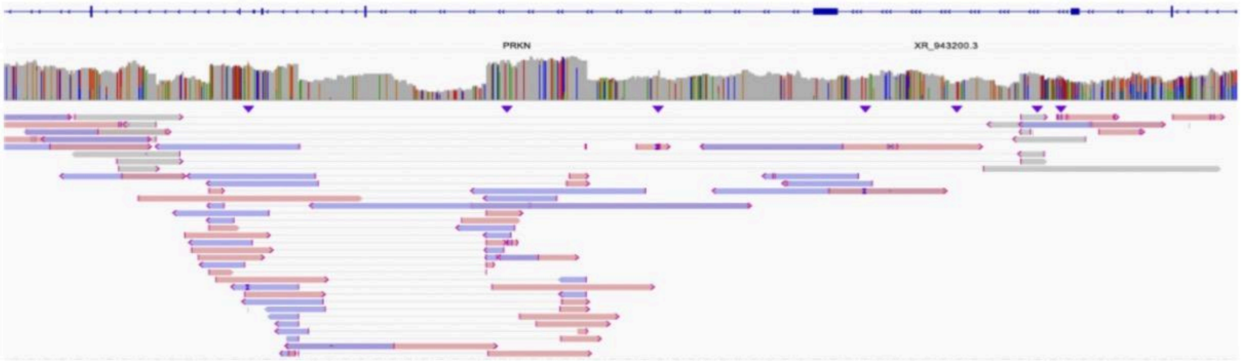

PRKN-11: Exon 2 deletion, Exon 3-4 duplication, and Exon 4 deletion

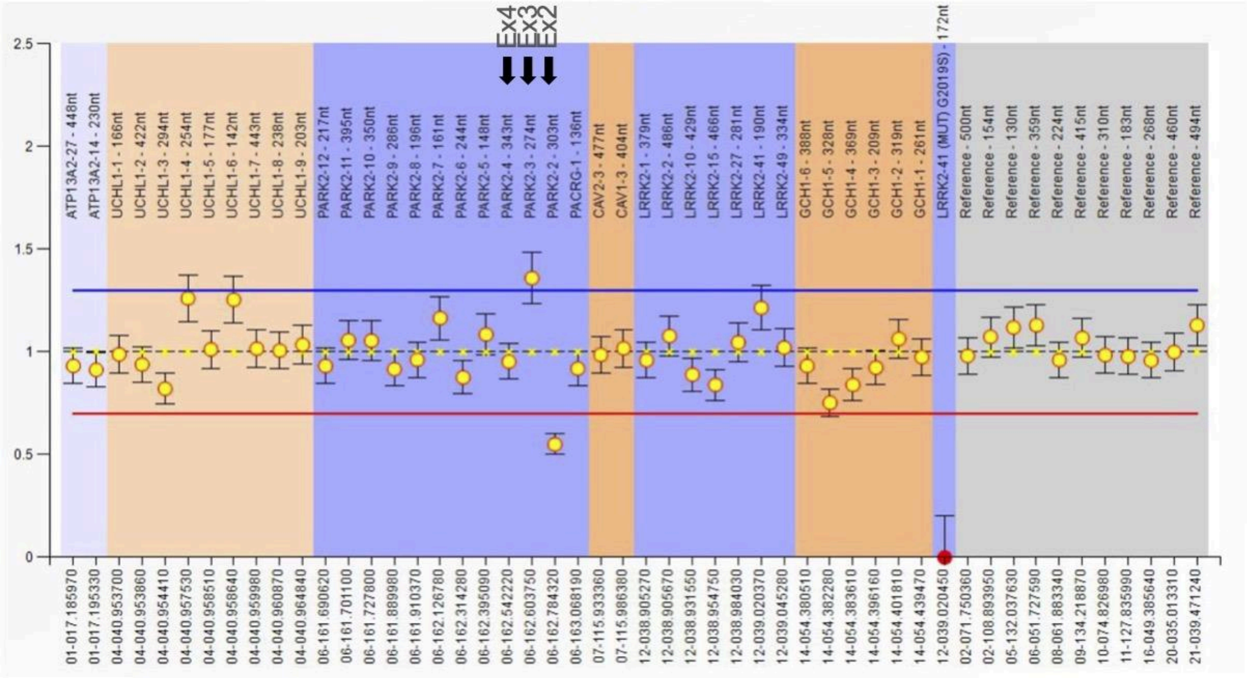


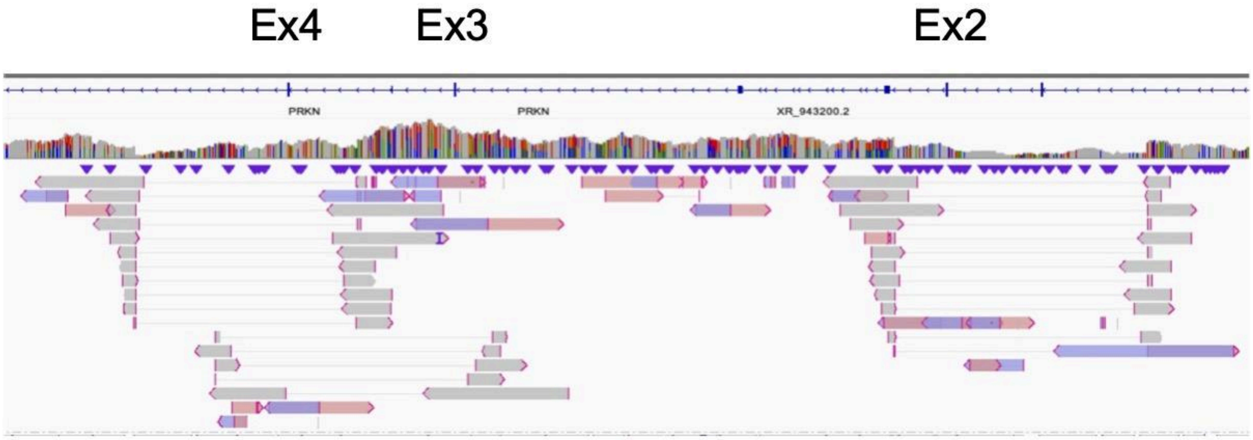

PRKN-12: Exon 5 duplication and Exon6 deletion

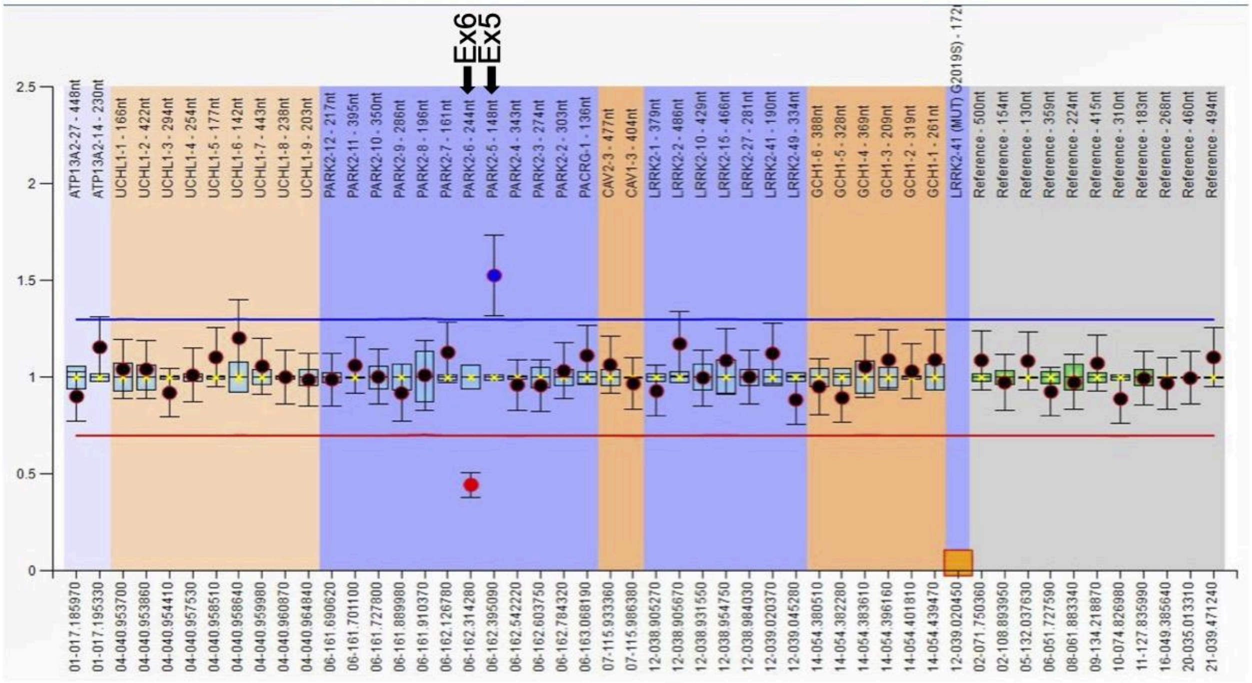

Ex6 Ex5

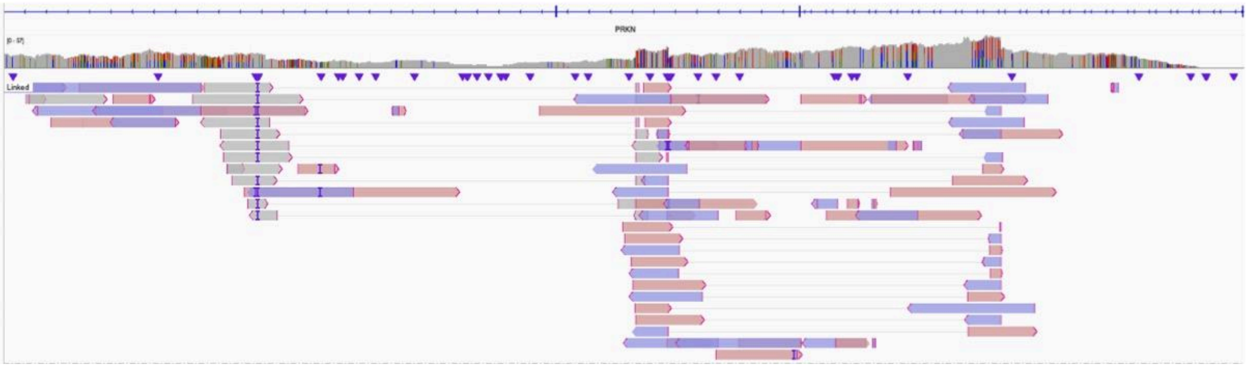

PRKN-13:

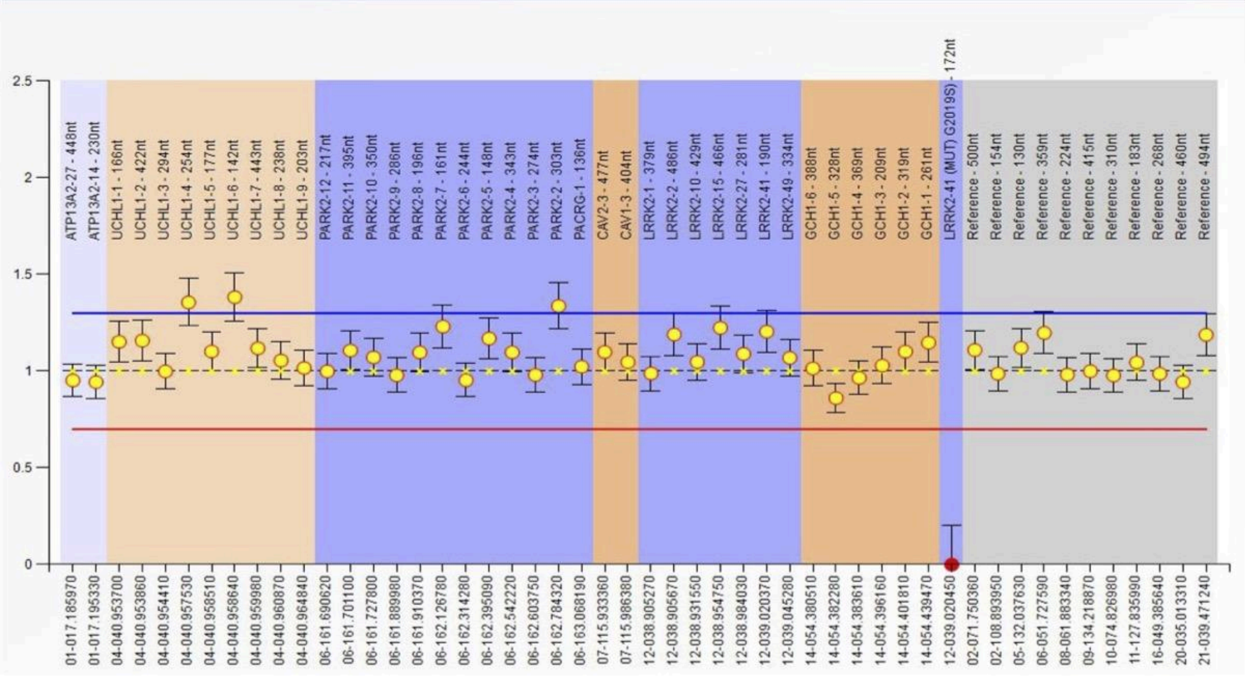
2.5
2
1.5
1
0.5
0
ATP13A2-27 - 448nt
ATP13A2-14 - 230nt
UCHL1-1 - 166nt
UCHL1-2 - 422nt
UCHL1-3 - 294nt
UCHL1-4 - 254nt
UCHL1-5 - 177nt
UCHL1-6 - 142nt
UCHL1-7 - 443nt
UCHL1-8 - 238nt
UCHL1-9 - 203nt
PARK2-12 - 217nt
PARK2-11 - 395nt
PARK2-10 - 350nt
PARK2-9 - 286nt
PARK2-8 - 196nt
PARK2-7 - 161nt
PARK2-6 - 244nt
PARK2-5 - 148nt
PARK2-4 - 343nt
PARK2-3 - 274nt
PARK2-2 - 303nt
PACRG-1 - 136nt
CAV2-3 - 477nt
CAV1-3 - 404nt
LRRK2-1 - 379nt
LRRK2-2 - 486nt
LRRK2-10 - 429nt
LRRK2-15 - 466nt
LRRK2-27 - 281nt
LRRK2-41 - 190nt
LRRK2-49 - 334nt
GCH1-6 - 388nt
GCH1-5 - 328nt
GCH1-4 - 369nt
GCH1-3 - 209nt
GCH1-2 - 319nt
GCH1-1 - 261nt
LRRK2-41 (MUT) G2019S) - 172nt
Reference - 500nt
Reference - 154nt
Reference - 130nt
Reference - 359nt
Reference - 224nt
Reference - 415nt
Reference - 310nt
Reference - 183nt
Reference - 268nt
Reference - 460nt
Reference - 494nt
01-017.185970
01-017.195330
04-040.953700
04-040.953860
04-040.954410
04-040.957530
04-040.958510
04-040.958640
04-040.959980
04-040.960870
04-040.964840
06-161.690620
06-161.701100
06-161.727800
06-161.889980
06-161.910370
06-162.126780
06-162.314280
06-162.395090
06-162.542220
06-162.603750
06-162.784320
06-163.068190
07-115.933360
07-115.986380
12-038.905270
12-038.905670
12-038.931550
12-038.954750
12-038.984030
12-039.020370
12-039.045280
14-054.380510
14-054.382280
14-054.383610
14-054.396160
14-054.401810
14-054.439470
12-039.020450
02-071.750360
02-108.893950
05-132.037630
06-051.727590
08-061.883340
09-134.218870
10-074.826980
11-127.835990
16-049.385640
20-035.013310
21-039.471240

PRKN-14:

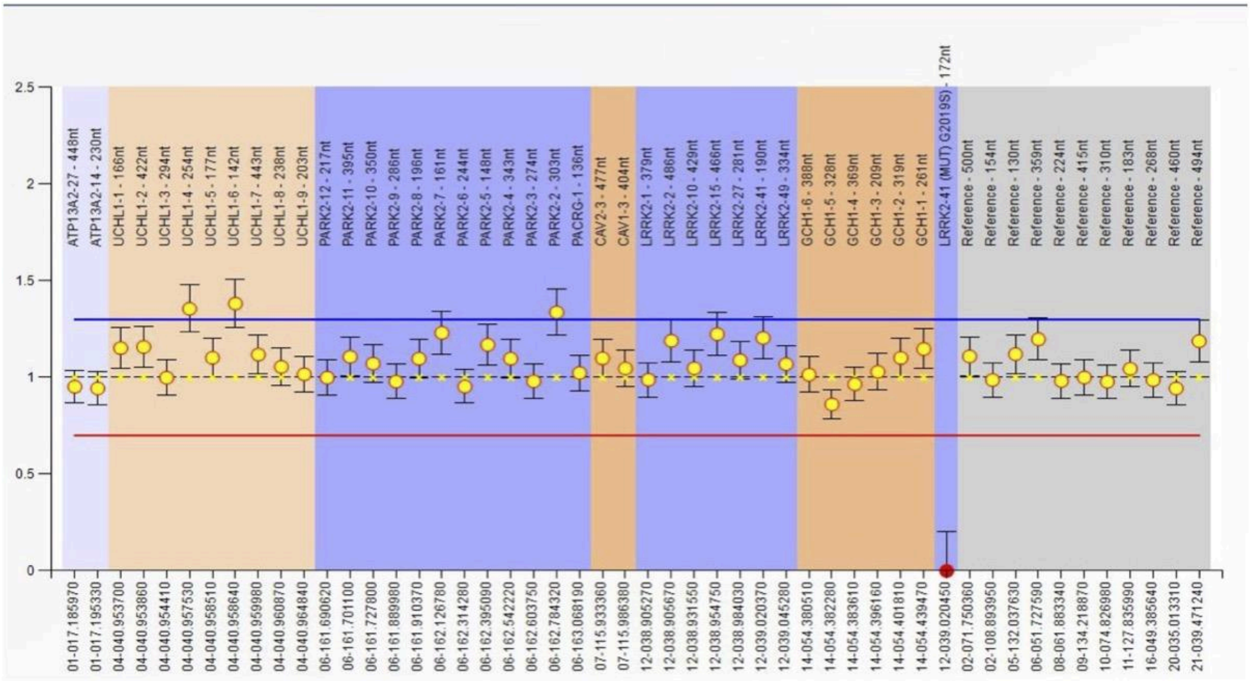
2.5
2
1.5
1
0.5
0
ATP13A2-27 - 448nt
ATP13A2-14 - 230nt
UCHL1-1 - 166nt
UCHL1-2 - 422nt
UCHL1-3 - 294nt
UCHL1-4 - 254nt
UCHL1-5 - 177nt
UCHL1-6 - 142nt
UCHL1-7 - 443nt
UCHL1-8 - 238nt
UCHL1-9 - 203nt
PARK2-12 - 217nt
PARK2-11 - 395nt
PARK2-10 - 350nt
PARK2-9 - 286nt
PARK2-8 - 196nt
PARK2-7 - 161nt
PARK2-6 - 244nt
PARK2-5 - 148nt
PARK2-4 - 343nt
PARK2-3 - 274nt
PARK2-2 - 303nt
PACRG-1 - 136nt
CAV2-3 - 477nt
CAV1-3 - 404nt
LRRK2-1 - 379nt
LRRK2-2 - 486nt
LRRK2-10 - 429nt
LRRK2-15 - 466nt
LRRK2-27 - 281nt
LRRK2-41 - 190nt
LRRK2-49 - 334nt
GCH1-6 - 388nt
GCH1-5 - 328nt
GCH1-4 - 369nt
GCH1-3 - 209nt
GCH1-2 - 319nt
GCH1-1 - 261nt
LRRK2-41 (MUT) G2019S) - 172nt
Reference - 500nt
Reference - 154nt
Reference - 130nt
Reference - 359nt
Reference - 224nt
Reference - 415nt
Reference - 310nt
Reference - 183nt
Reference - 268nt
Reference - 460nt
Reference - 494nt
01-017.185970
01-017.195330
04-040.953700
04-040.953860
04-040.954410
04-040.957530
04-040.958510
04-040.958640
04-040.959980
04-040.960870
04-040.964840
06-161.690620
06-161.701100
06-161.727800
06-161.889980
06-161.910370
06-162.126780
06-162.314280
06-162.395090
06-162.542220
06-162.603750
06-162.784320
06-163.068190
07-115.933360
07-115.986380
12-038.905270
12-038.905670
12-038.931550
12-038.954750
12-038.984030
12-039.020370
12-039.045280
14-054.380510
14-054.382280
14-054.383610
14-054.396160
14-054.401810
14-054.439470
12-039.020450
02-071.750360
02-108.893950
05-132.037630
06-051.727590
08-061.883340
09-134.218870
10-074.826980
11-127.835990
16-049.385640
20-035.013310
21-039.471240

PRKN-15: Exon 6-7 duplication

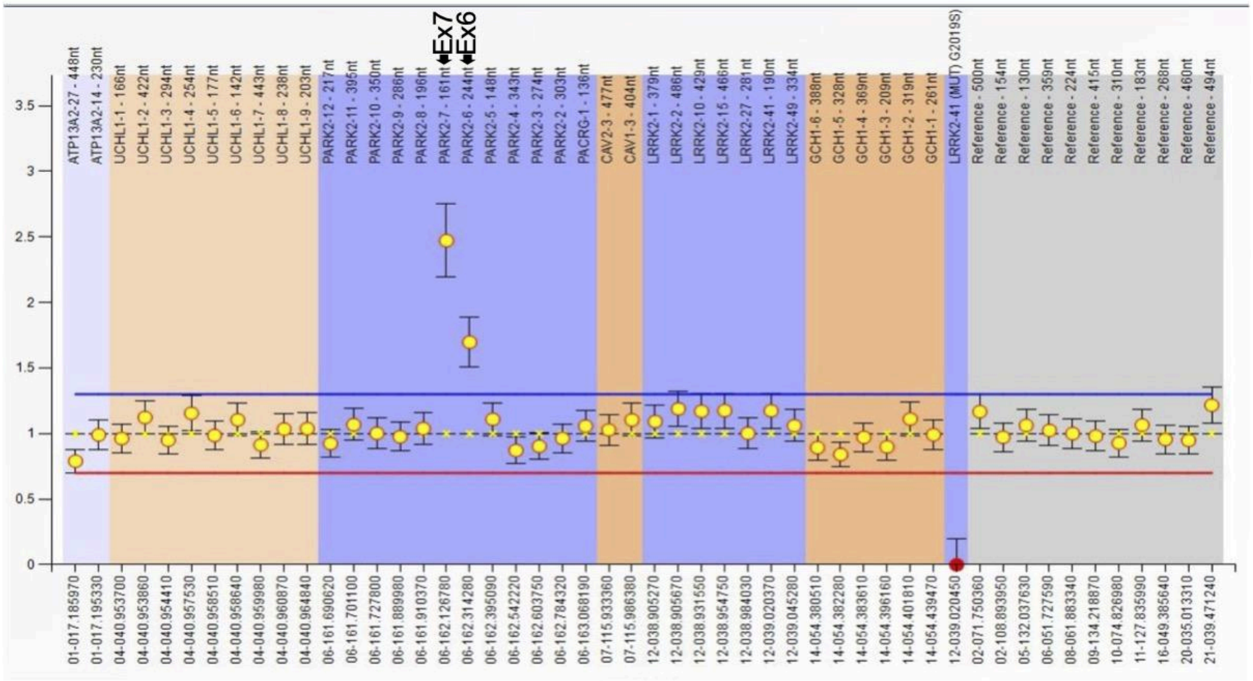


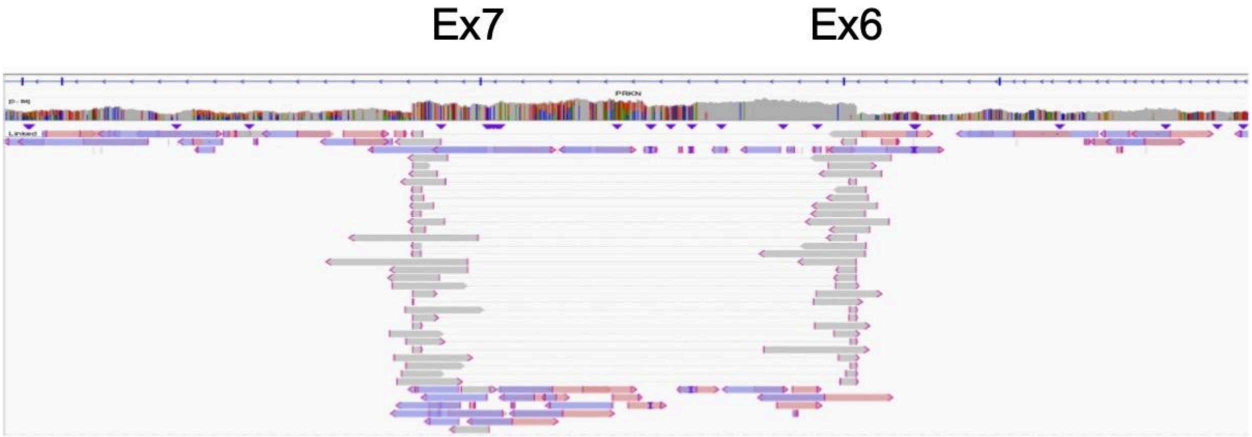

PRKN-16:

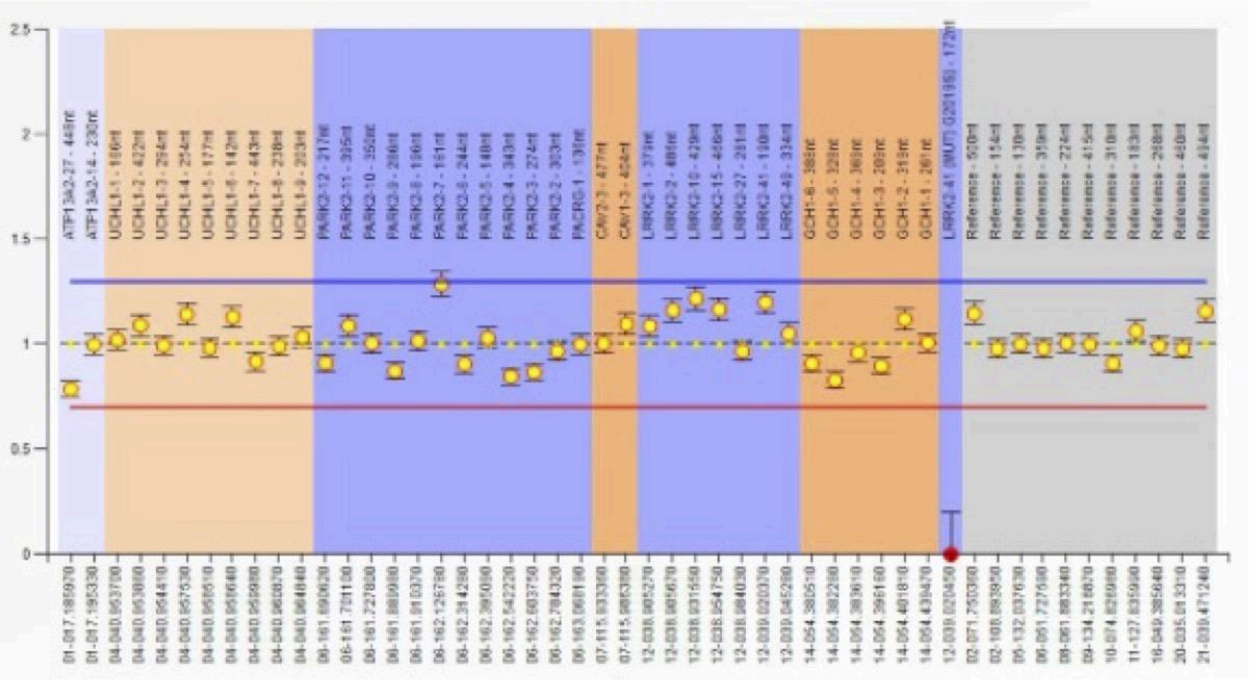

2.5
2
1.5
1
0.5
0
ATP13A2-27 - 448nt
ATP13A2-14 - 230nt
UCHL1-1 - 186nt
UCHL1-2 - 422nt
UCHL1-3 - 294nt
UCHL1-4 - 254nt
UCHL1-5 - 177nt
UCHL1-6 - 142nt
UCHL1-7 - 443nt
UCHL1-8 - 238nt
UCHL1-9 - 283nt
PARK2-12 - 217nt
PARK2-11 - 395nt
PARK2-10 - 352nt
PARK2-9 - 286nt
PARK2-8 - 156nt
PARK2-7 - 161nt
PARK2-6 - 244nt
PARK2-5 - 148nt
PARK2-4 - 343nt
PARK2-3 - 274nt
PARK2-2 - 303nt
PACRG-1 - 136nt
CAV2-3 - 477nt
CAV1-3 - 454nt
LRRK2-1 - 373nt
LRRK2-2 - 481nt
LRRK2-10 - 429nt
LRRK2-15 - 456nt
LRRK2-27 - 281nt
LRRK2-41 - 190nt
LRRK2-49 - 334nt
GCH1-6 - 388nt
GCH1-5 - 328nt
GCH1-4 - 369nt
GCH1-3 - 209nt
GCH1-2 - 319nt
GCH1-1 - 261nt
LRRK2-41 (MUT) G2019S - 172nt
Reference - 500nt
Reference - 154nt
Reference - 130nt
Reference - 359nt
Reference - 224nt
Reference - 415nt
Reference - 310nt
Reference - 183nt
Reference - 288nt
Reference - 490nt
Reference - 494nt

PRKN-17: Exon 1 deletion and Exon 7 deletion

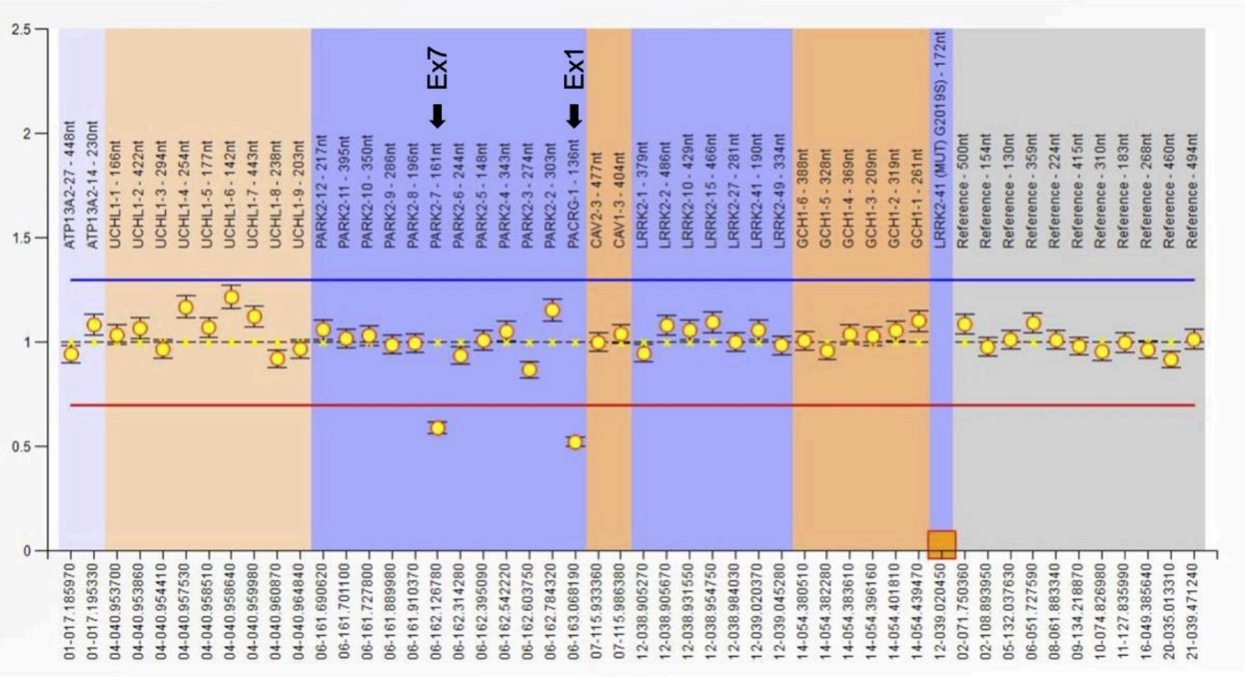


Ex1

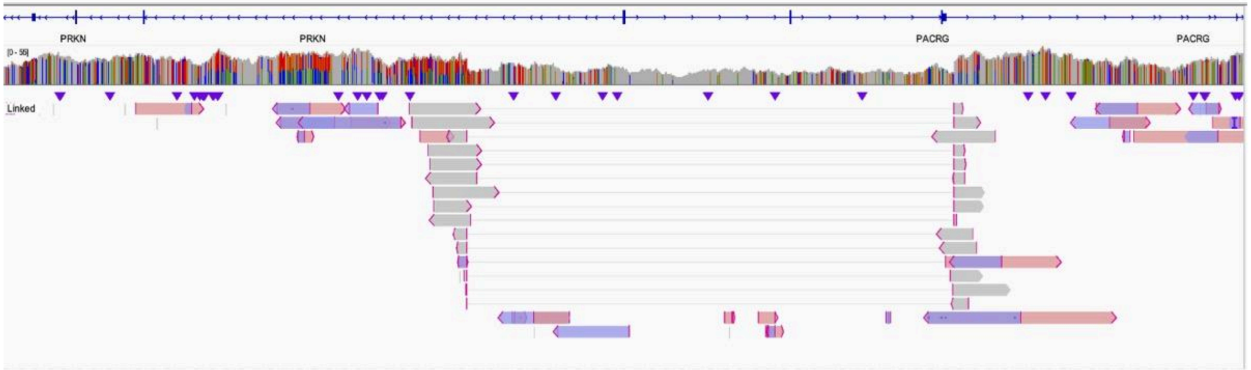


Ex7

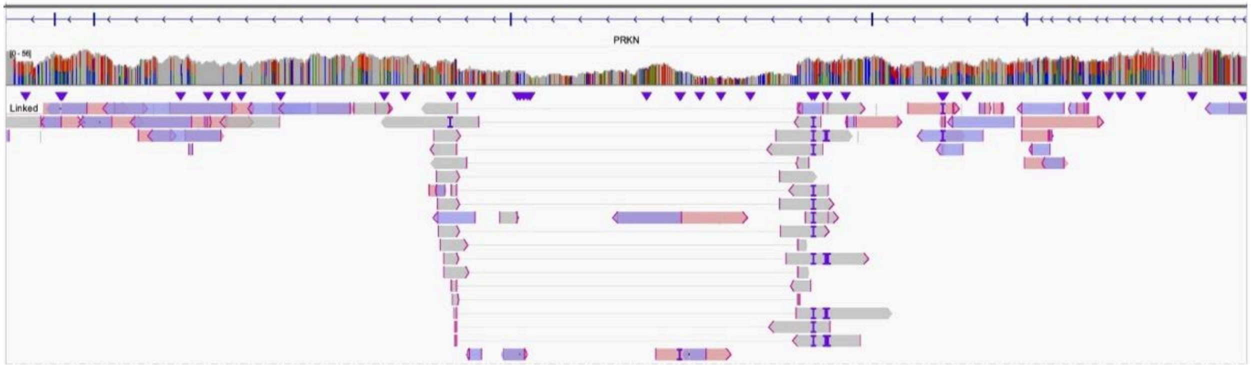

PRKN-18: Exon 2-4 duplication and Exon 3-5 deletion

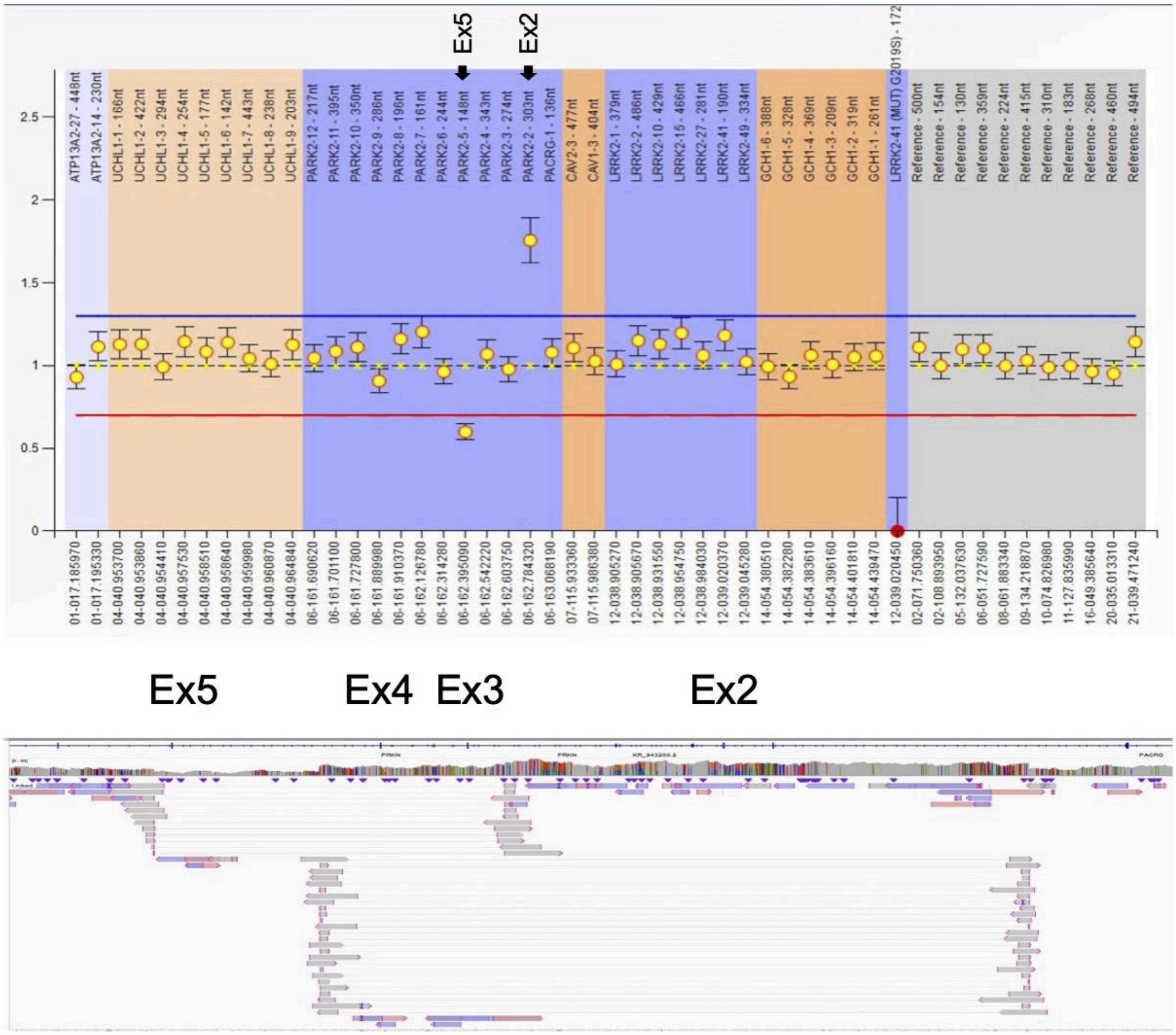

PRKN-19: Exon 2 duplication

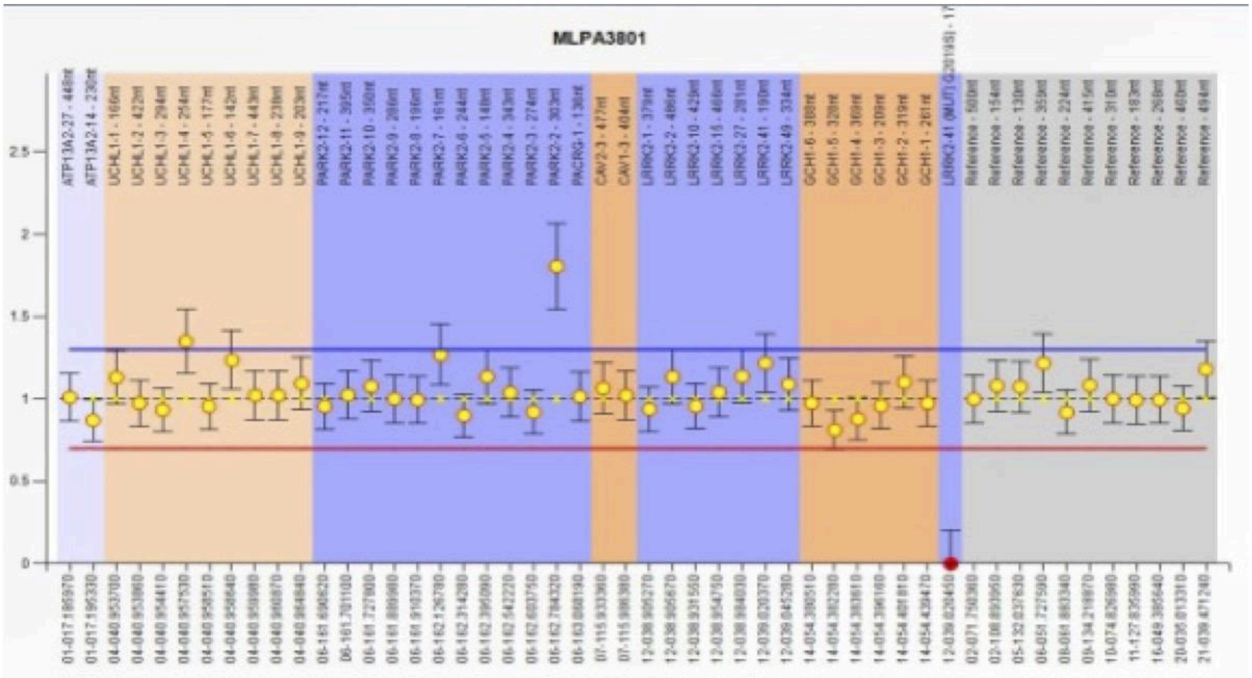

PRKN-20: Exon 4 deletion

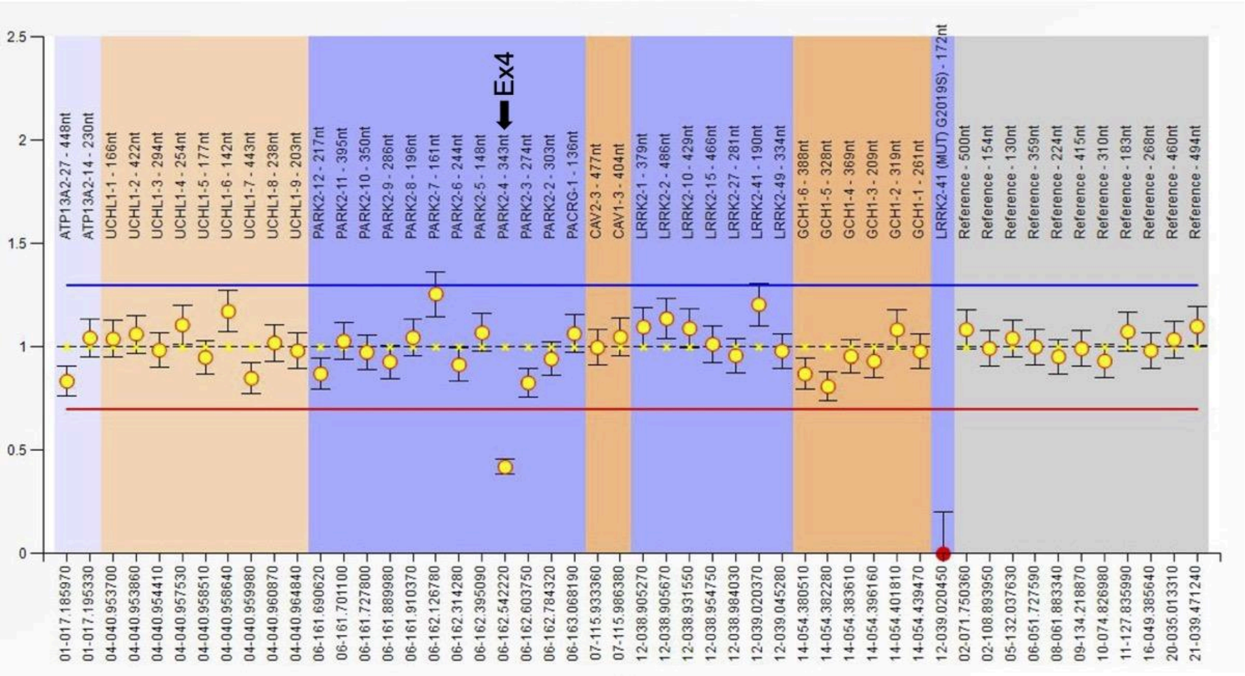


## Ex4

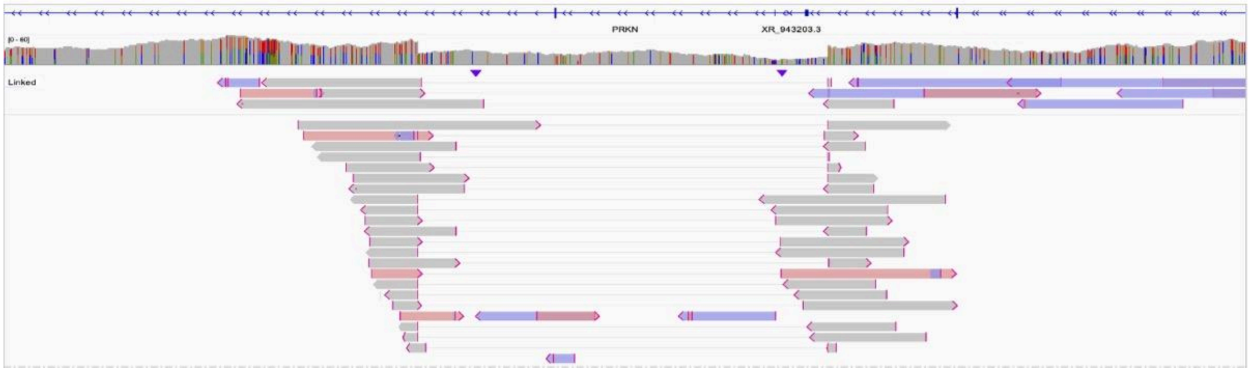

PRKN-21: Exon 6 duplication

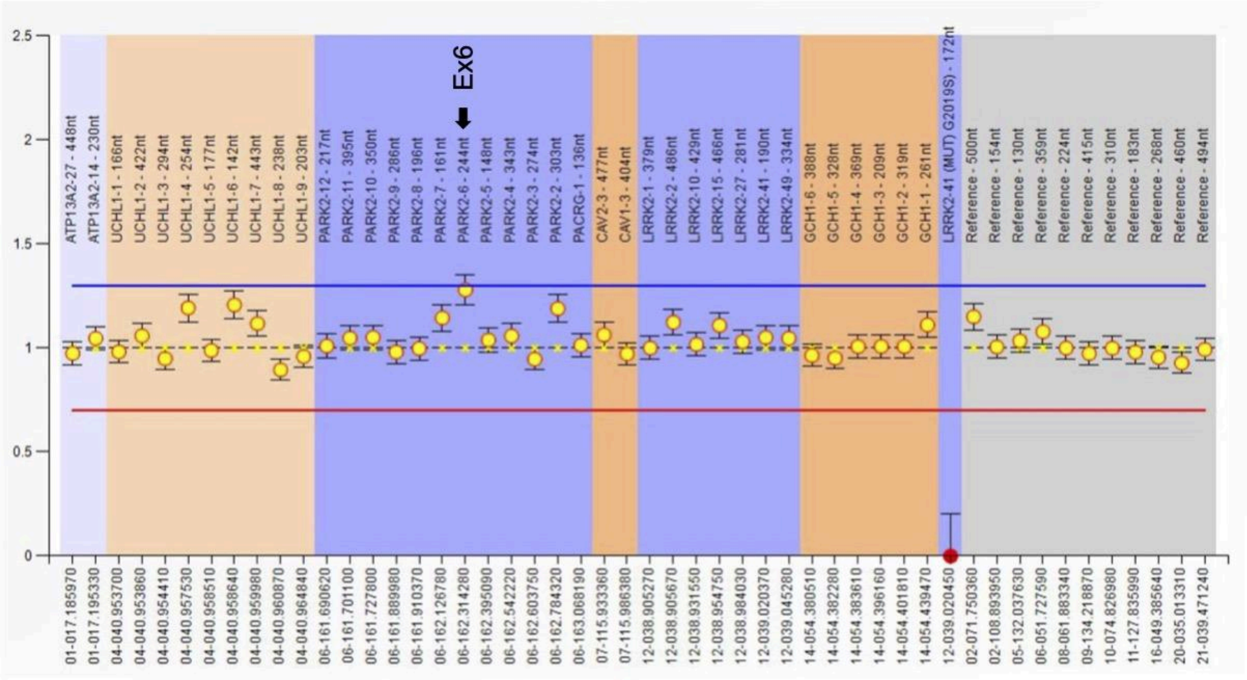


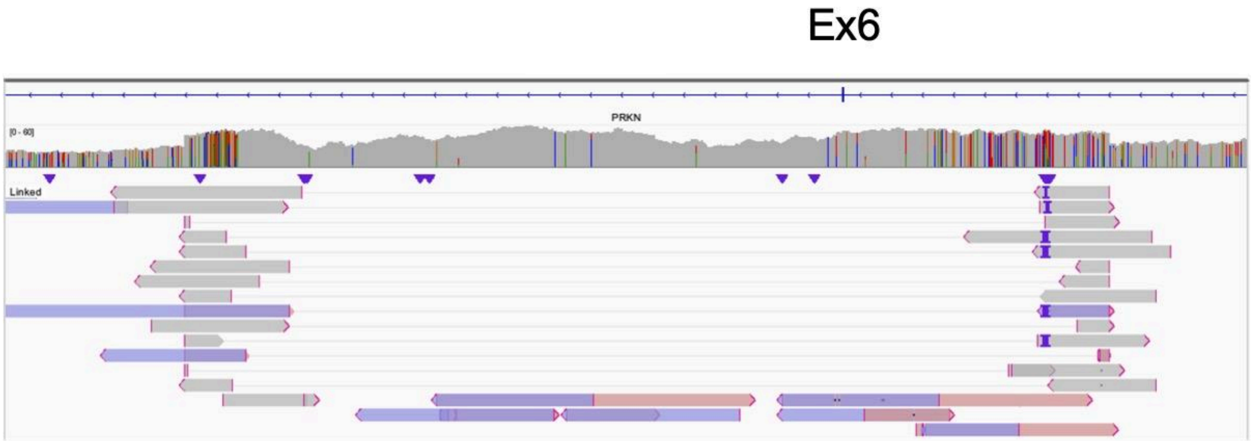

PRKN-22:

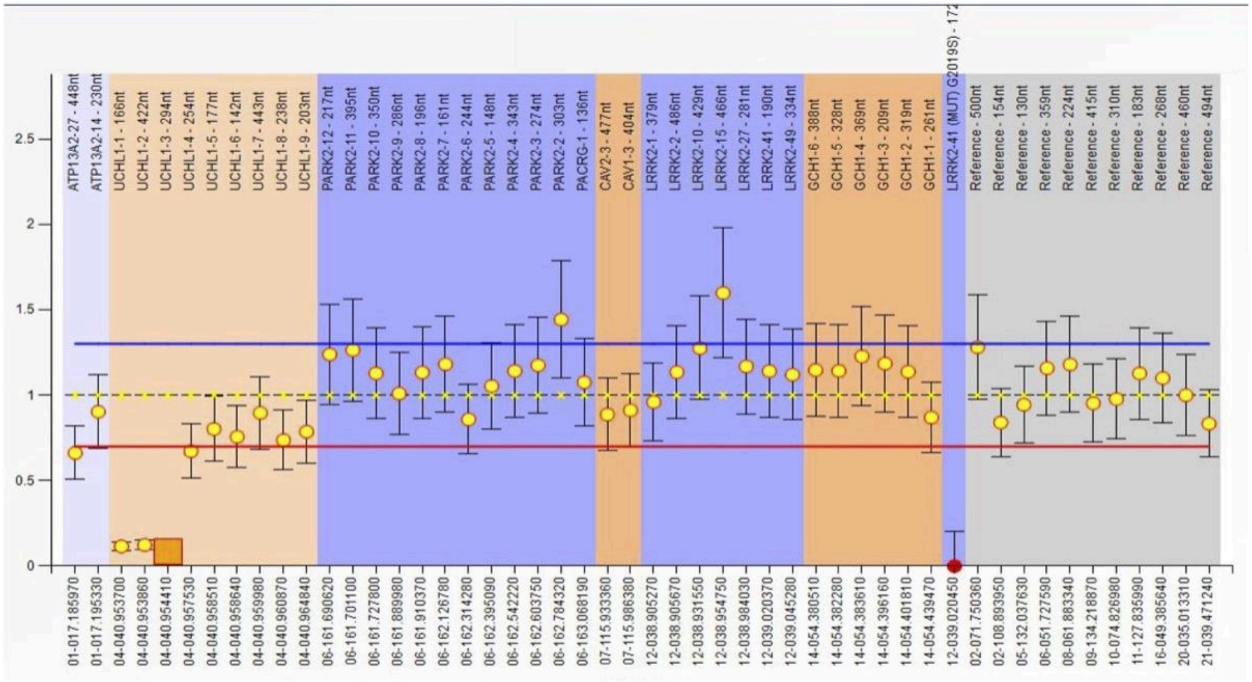
ATP13A2-27 - 448nt
ATP13A2-14 - 230nt
UCHL1-1 - 166nt
UCHL1-2 - 422nt
UCHL1-3 - 294nt
UCHL1-4 - 254nt
UCHL1-5 - 177nt
UCHL1-6 - 142nt
UCHL1-7 - 443nt
UCHL1-8 - 238nt
UCHL1-9 - 203nt
PARK2-12 - 217nt
PARK2-11 - 395nt
PARK2-10 - 350nt
PARK2-9 - 286nt
PARK2-8 - 196nt
PARK2-7 - 161nt
PARK2-6 - 244nt
PARK2-5 - 148nt
PARK2-4 - 343nt
PARK2-3 - 274nt
PARK2-2 - 303nt
PACRG-1 - 136nt
CAV2-3 - 477nt
CAV1-3 - 404nt
LRRK2-1 - 379nt
LRRK2-2 - 486nt
LRRK2-10 - 429nt
LRRK2-15 - 466nt
LRRK2-27 - 281nt
LRRK2-41 - 190nt
LRRK2-49 - 334nt
GCH1-6 - 388nt
GCH1-5 - 328nt
GCH1-4 - 369nt
GCH1-3 - 209nt
GCH1-2 - 319nt
GCH1-1 - 261nt
LRRK2-41 (MUT) G2019S) - 172
Reference - 500nt
Reference - 154nt
Reference - 130nt
Reference - 359nt
Reference - 224nt
Reference - 415nt
Reference - 310nt
Reference - 183nt
Reference - 268nt
Reference - 460nt
Reference - 494nt
2.5
2
1.5
1
0.5
0
01-017.185970
01-017.195330
04-040.953700
04-040.953860
04-040.954410
04-040.957530
04-040.958510
04-040.958640
04-040.959980
04-040.960870
04-040.964840
06-161.690620
06-161.701100
06-161.727800
06-161.889980
06-161.910370
06-162.126780
06-162.314280
06-162.395090
06-162.542220
06-162.603750
06-162.784320
06-163.068190
07-115.933360
07-115.986380
12-038.905270
12-038.905670
12-038.931550
12-038.954750
12-038.984030
12-039.020370
12-039.045280
14-054.380510
14-054.382280
14-054.383610
14-054.396160
14-054.401810
14-054.439470
12-039.020450
02-071.750360
02-108.893950
05-132.037630
06-051.727590
08-061.883340
09-134.218870
10-074.826980
11-127.835990
16-049.385640
20-035.013310
21-039.471240

PRKN-23: Exon 3-4 deletion
No long-read sequencing data due to the DNA quality.

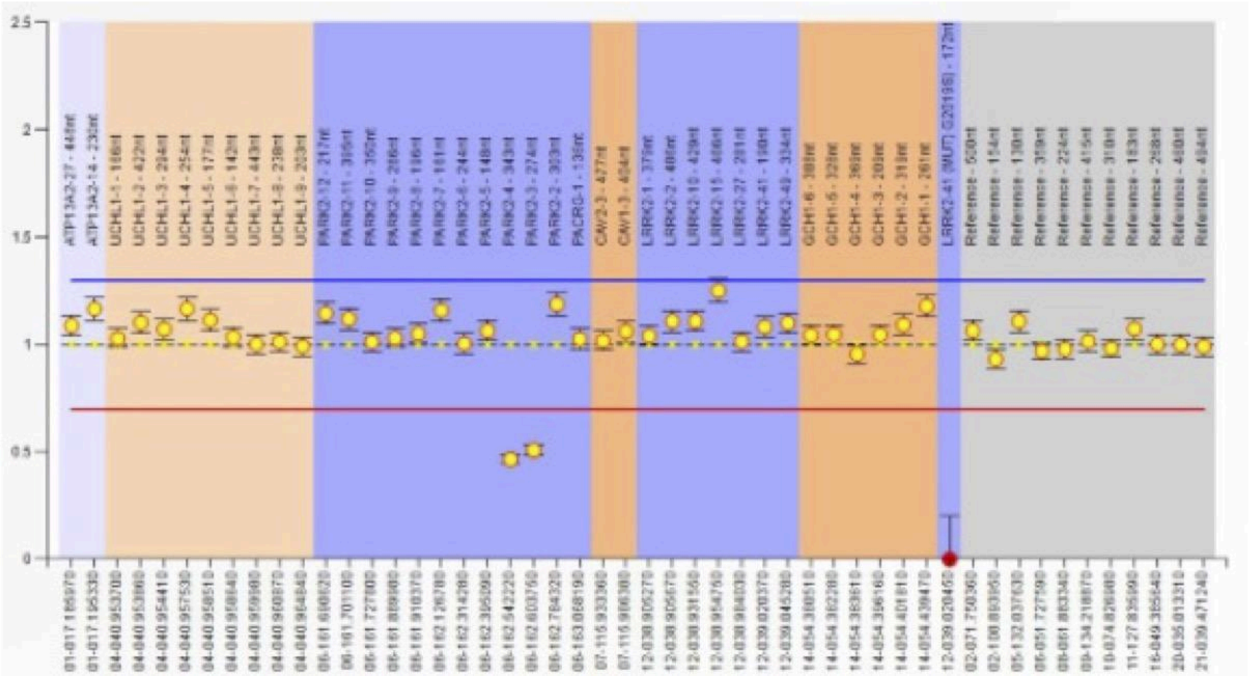

PRKN-24: Exon 2-4 deletion and Exon 6 duplication

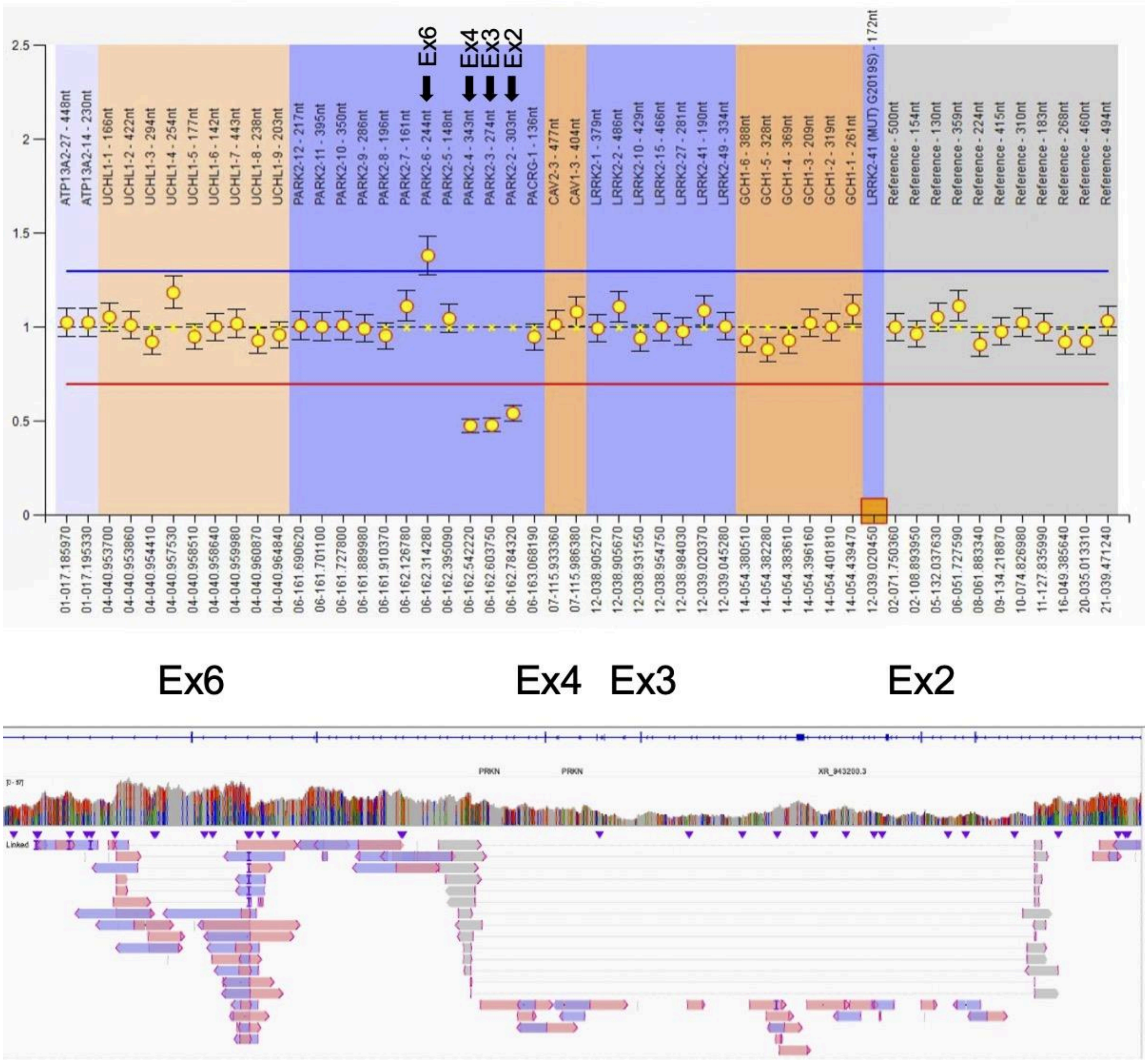

PRKN-25: Exon 5-6 duplication and Exon 6 duplication

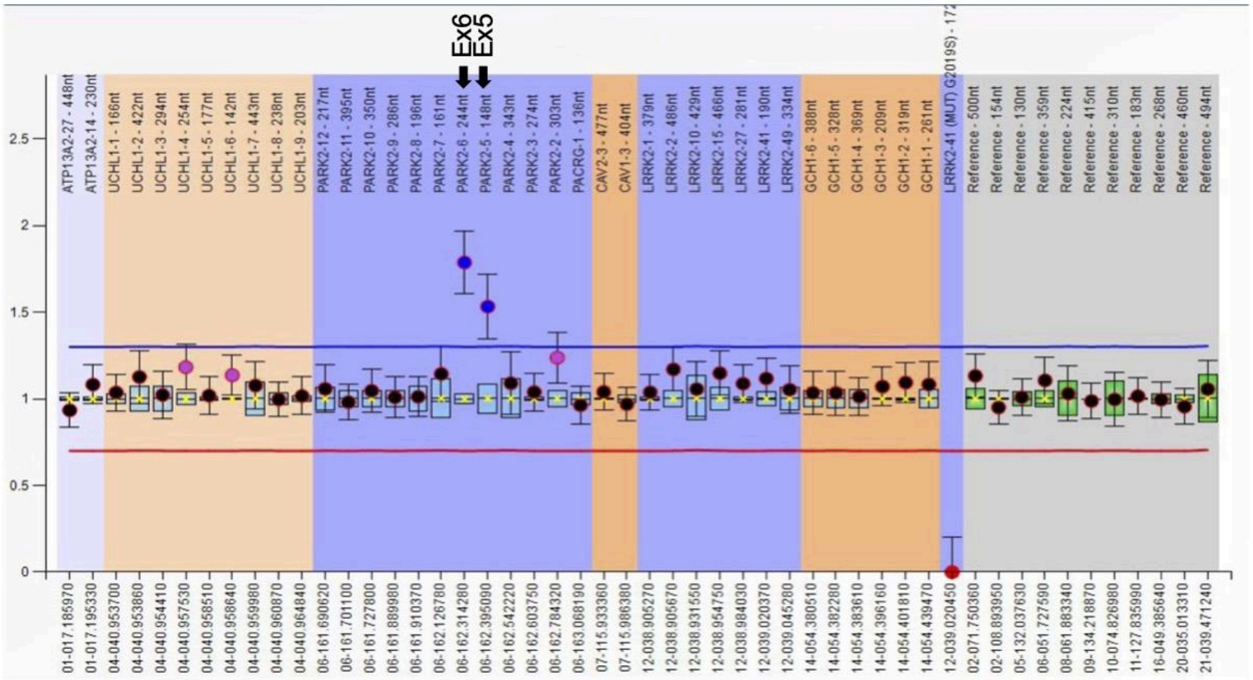


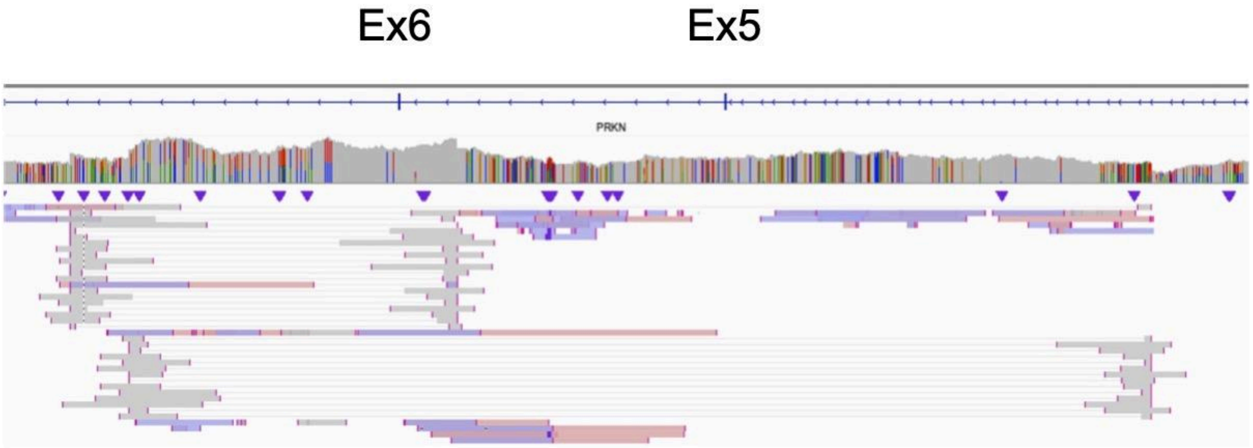

PRKN-26: Exon 2-4 deletion and Exon 7 deletion

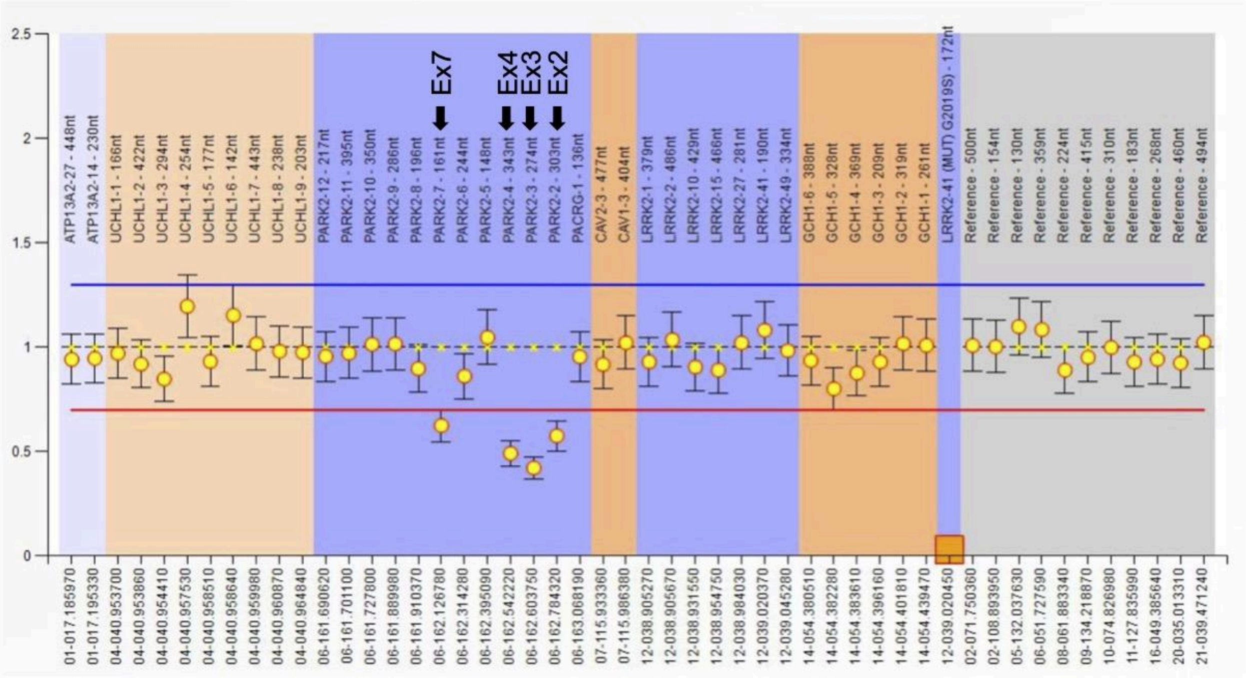


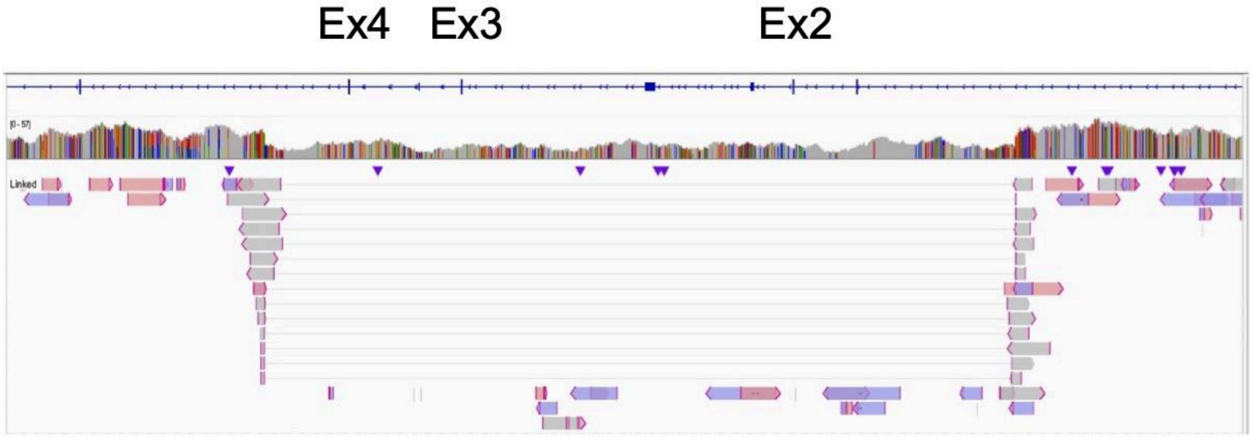


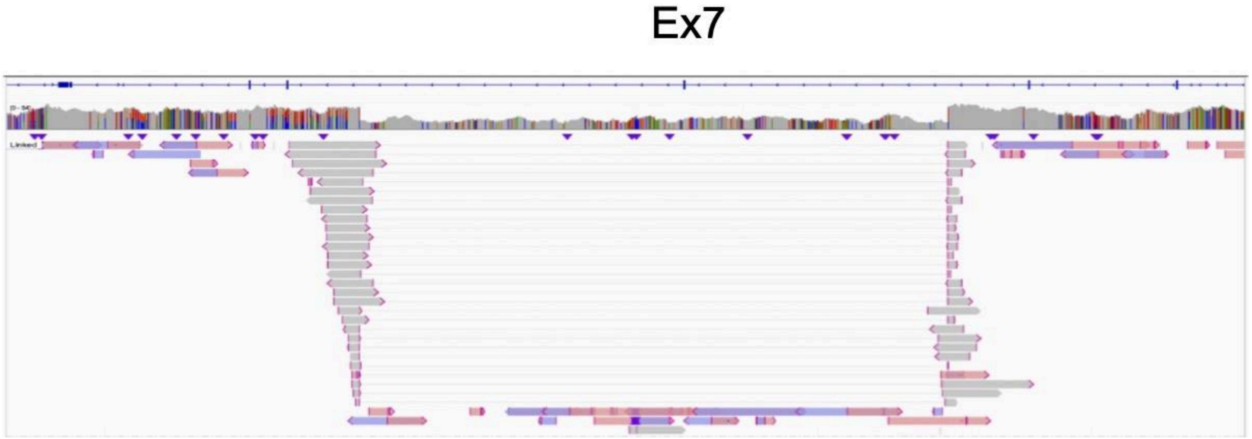

PRKN-27: Exon 3-4 deletion and Exon 5 duplication

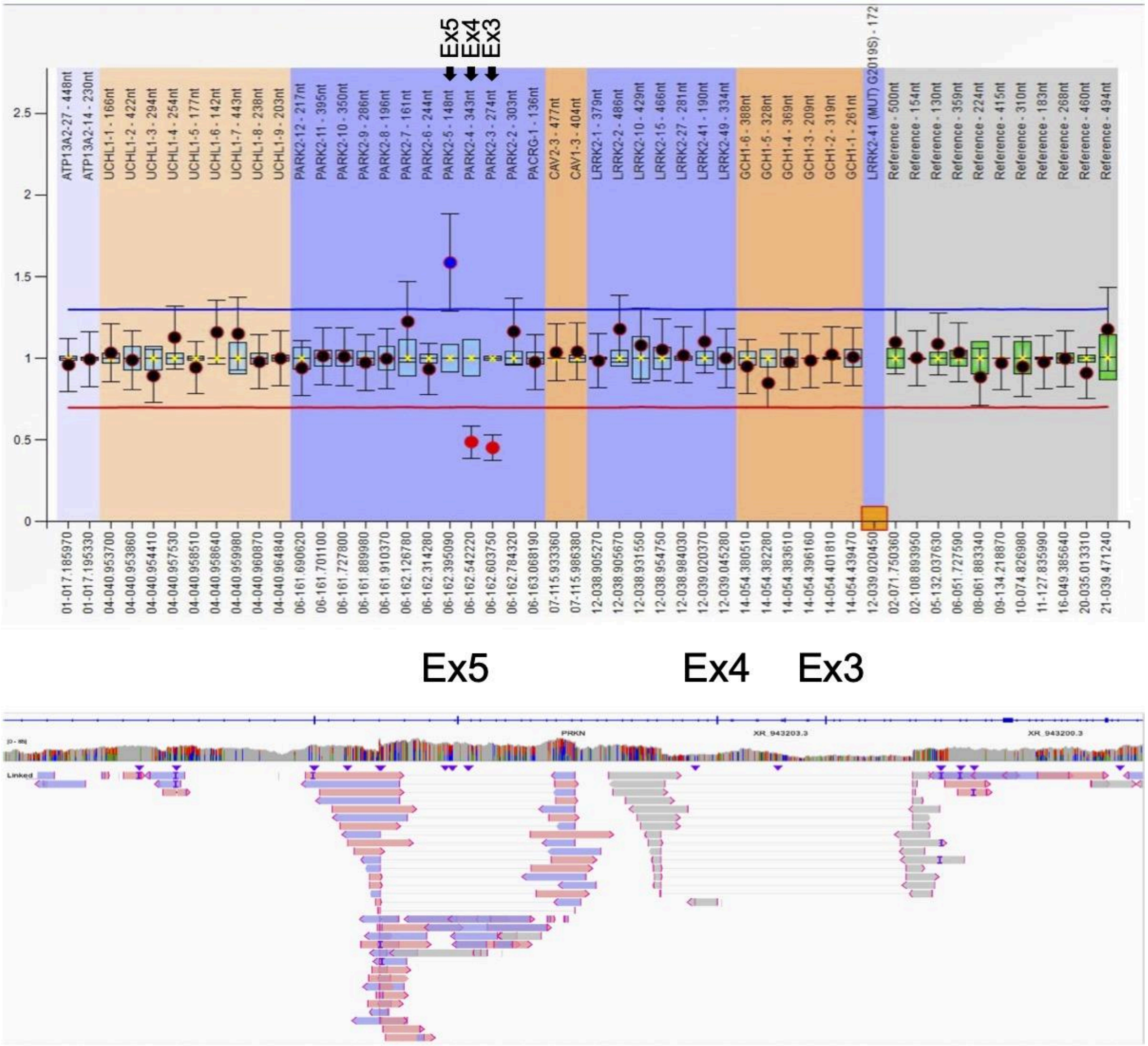

PRKN-28: Exon 3-4 deletion

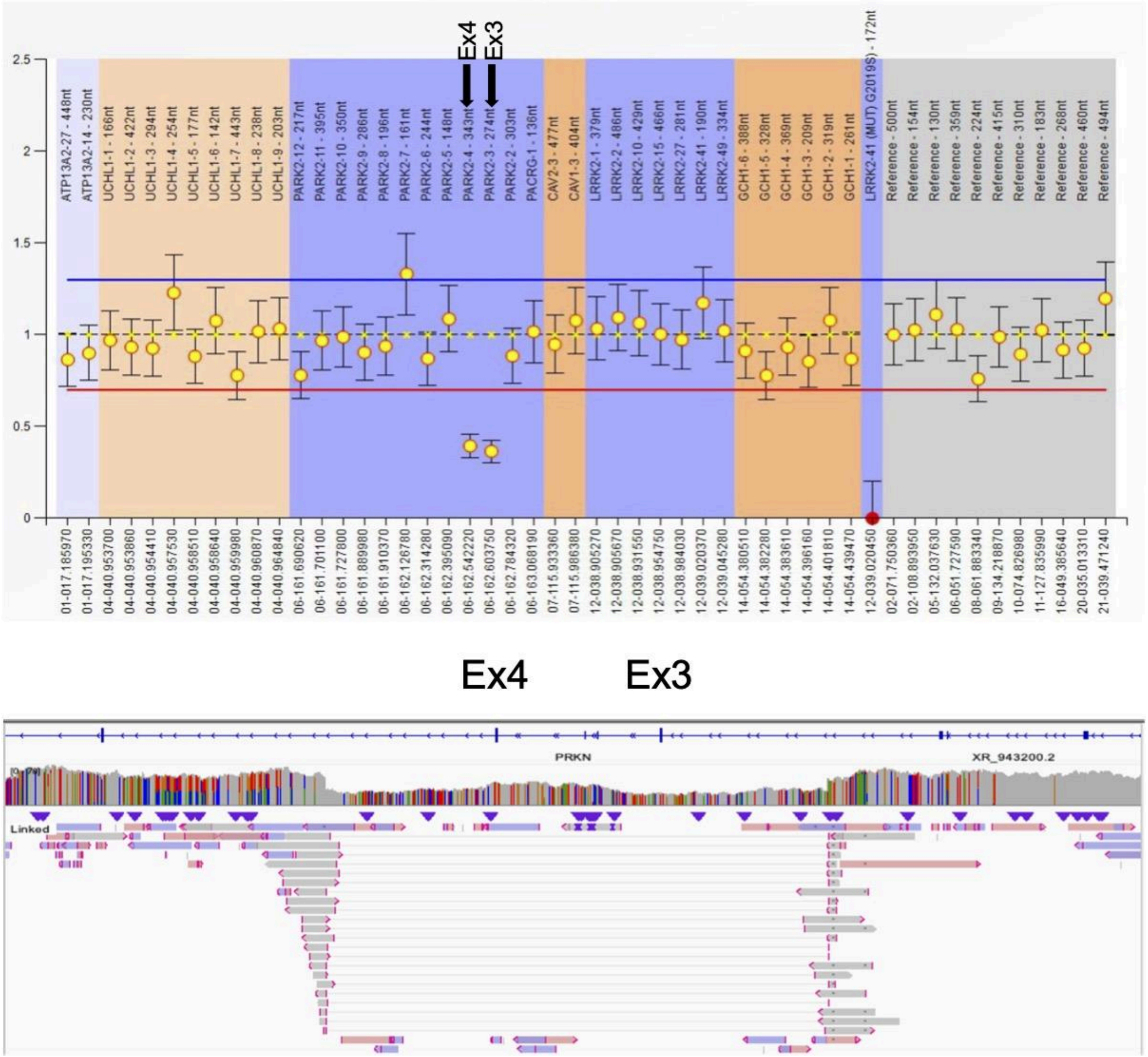

PRKN-29: Exon 3-7 duplication

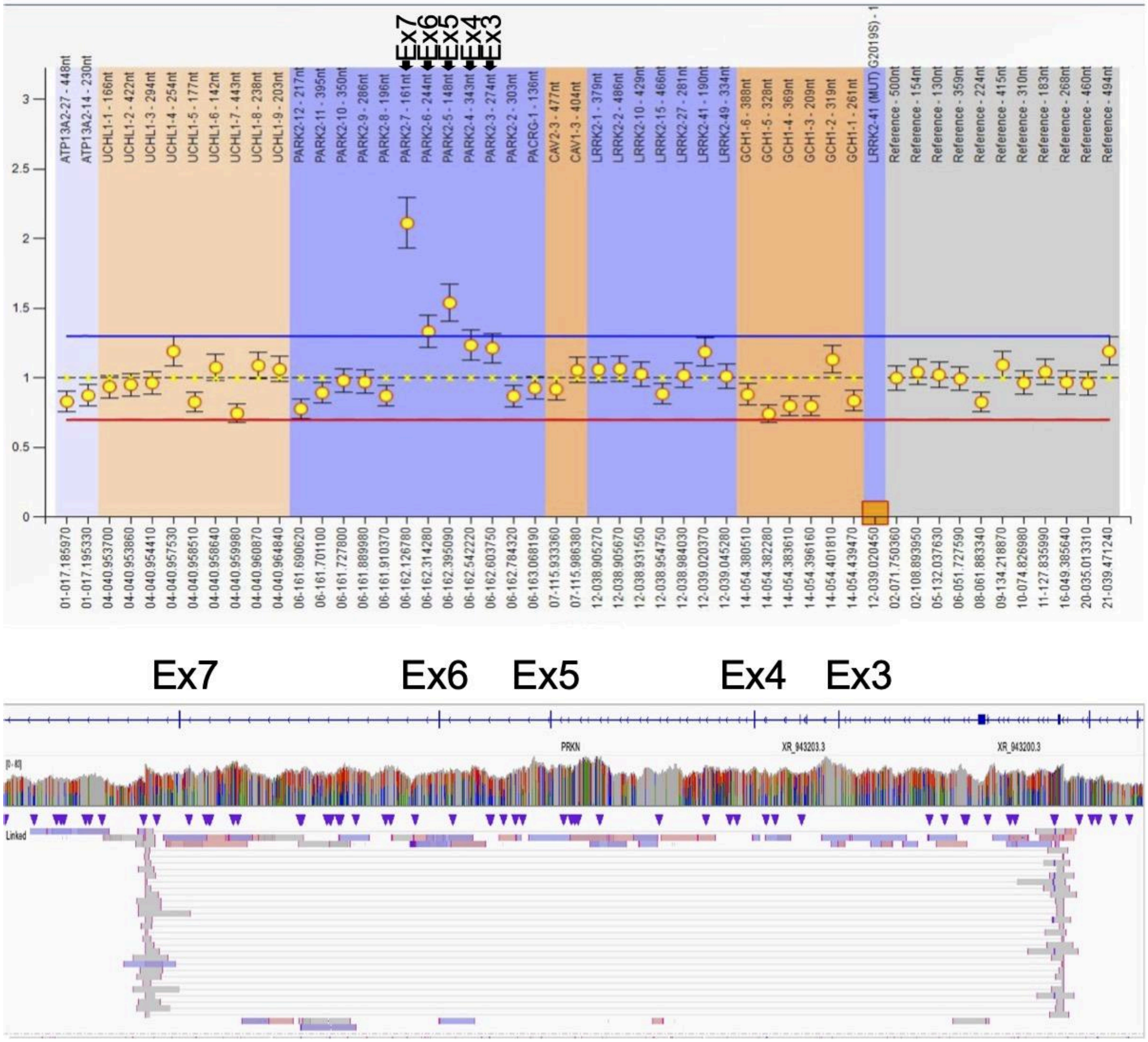

PRKN-30: Exon 3 deletion
The result of MLPA for exon 7 was judged as normal combined with qPCR result.

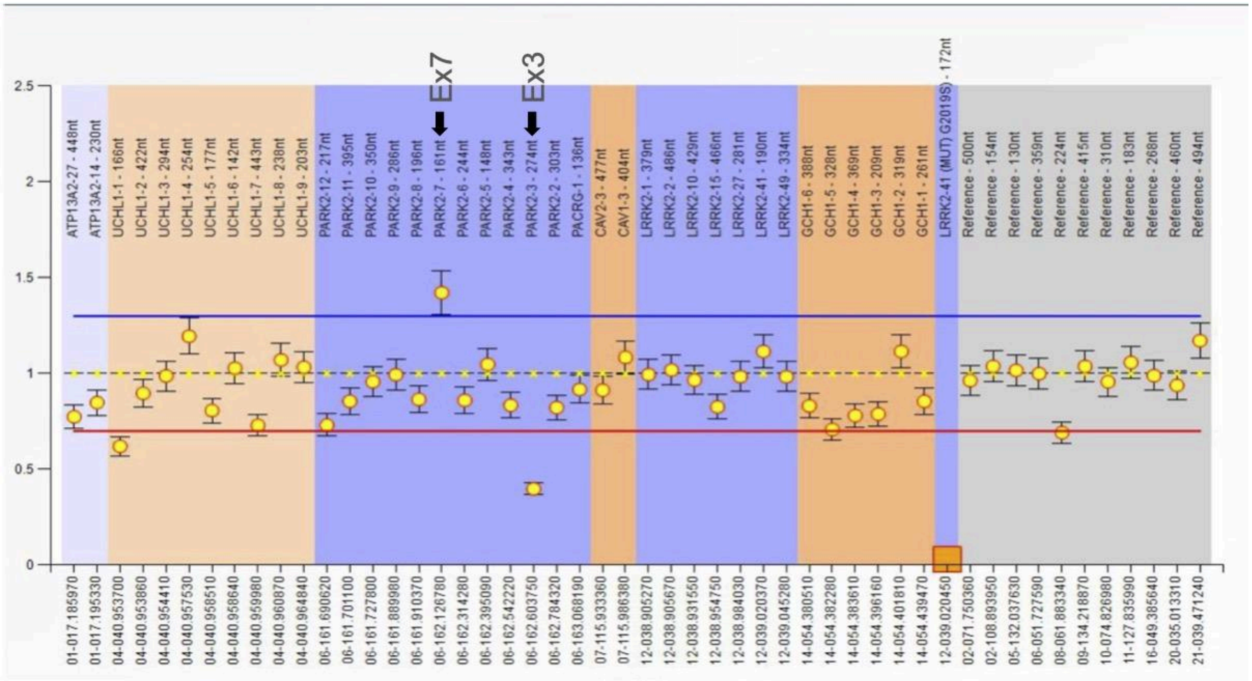


Ex3

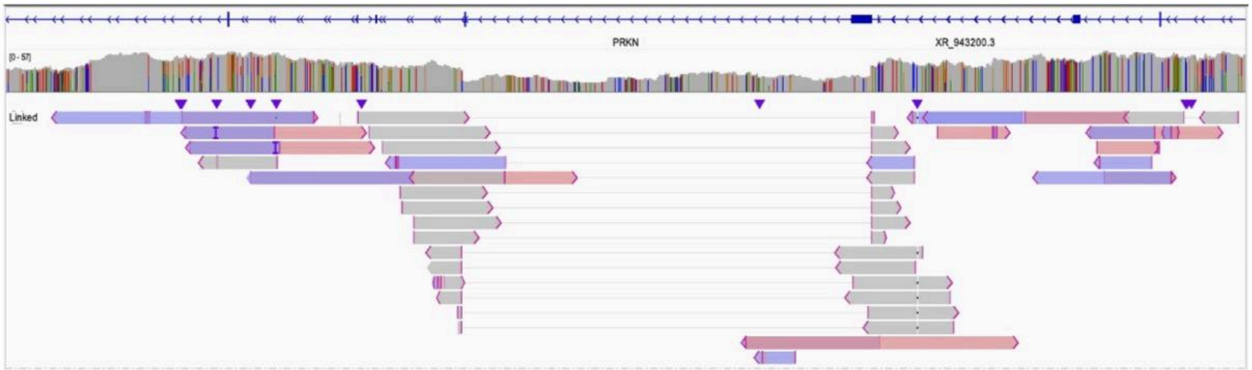


Ex7

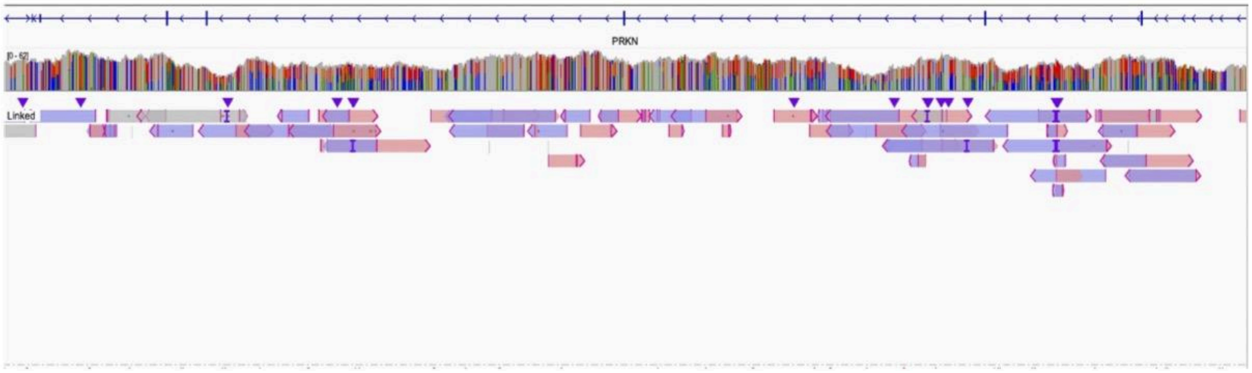

PRKN-31: Exon 3-4 deletion

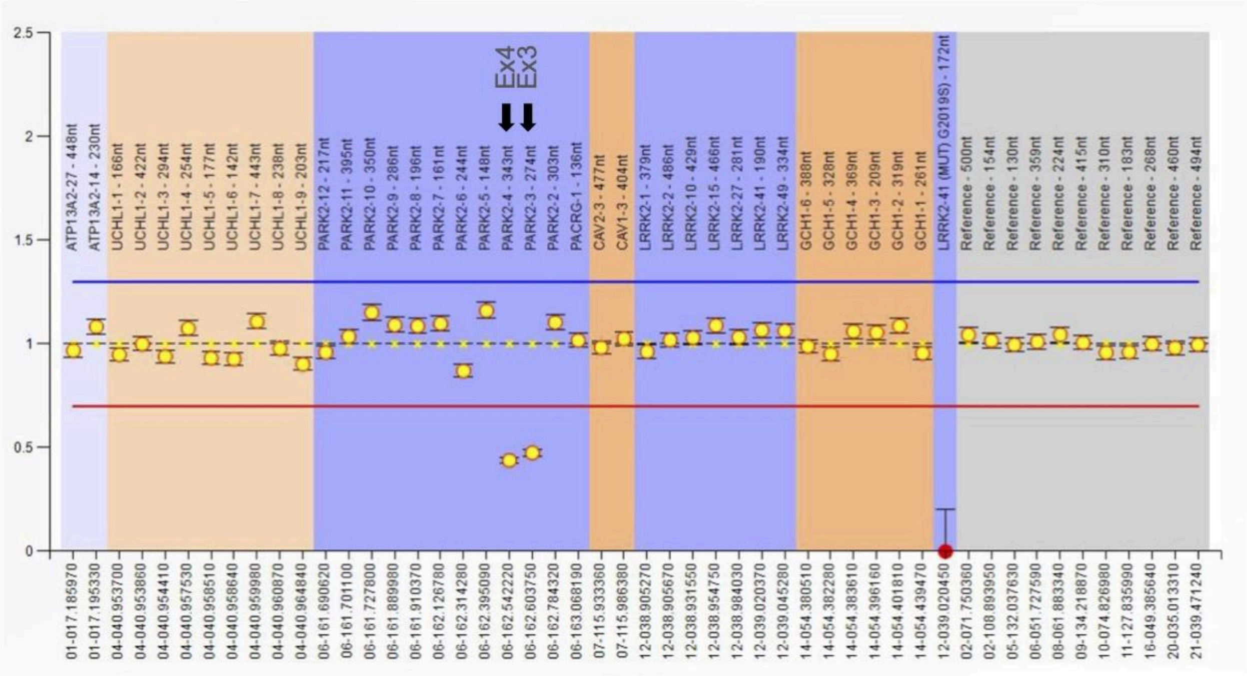


Ex4 Ex3

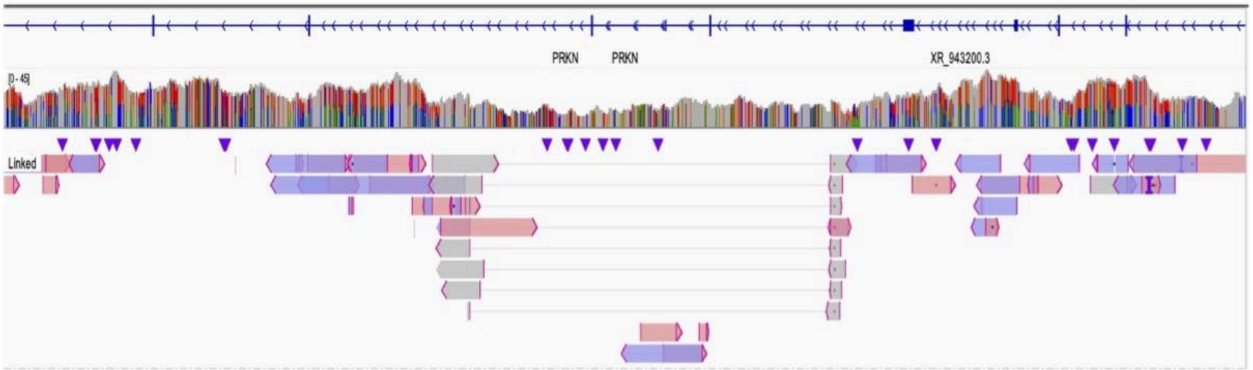

PRKN-32: Exon 2 deletion and Exon 3 deletion

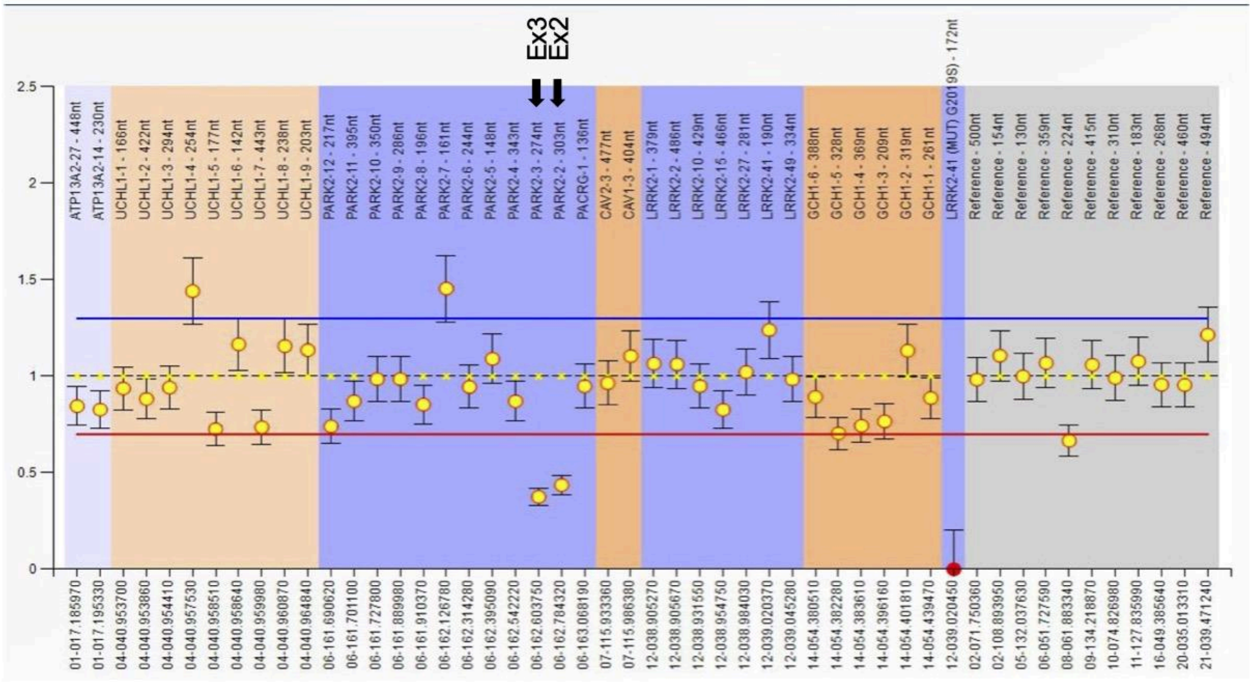


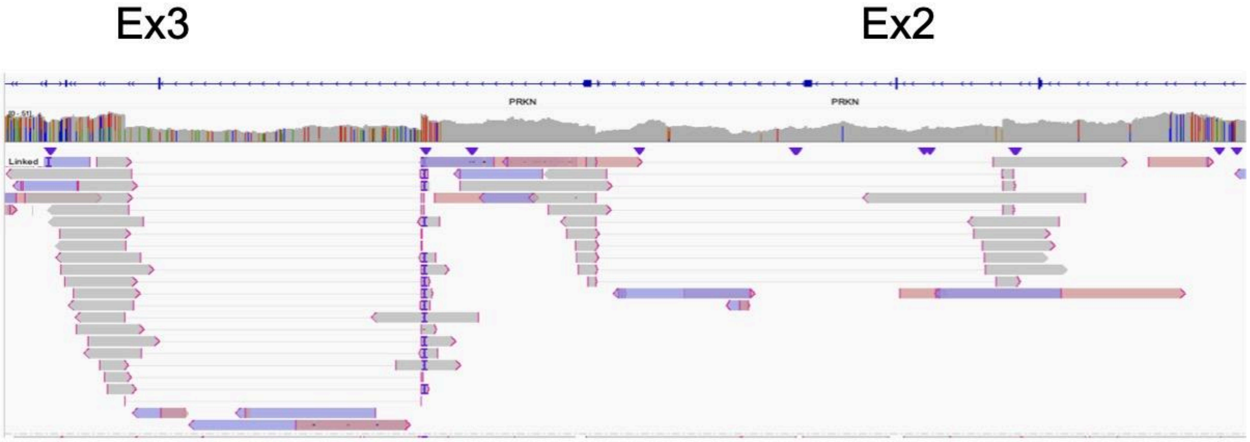

PRKN-33: Exon 3 deletion

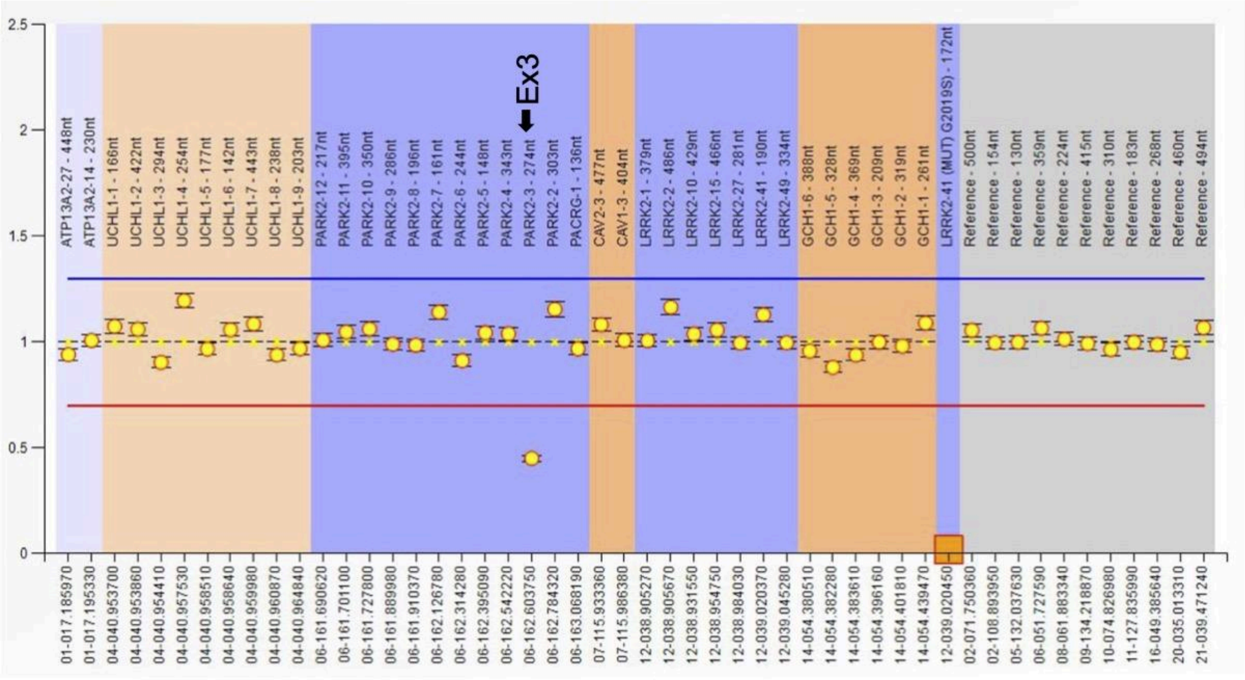


Ex3

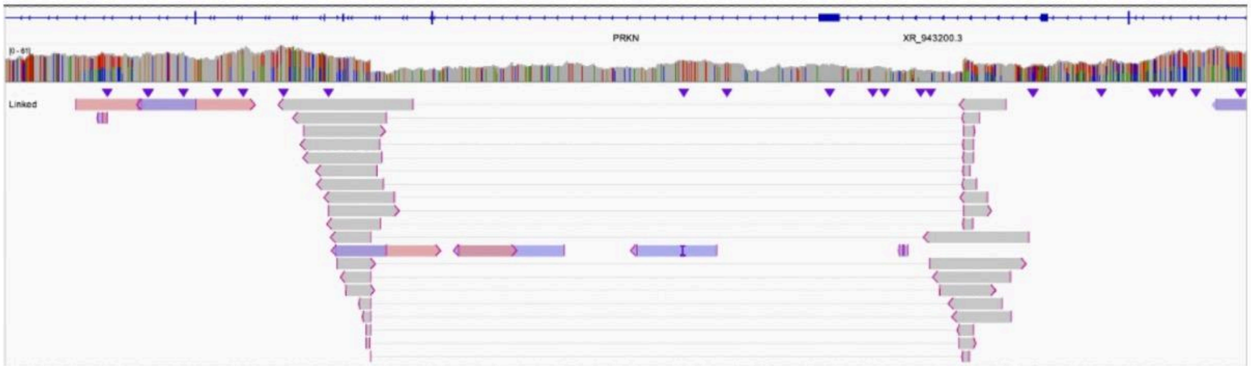

PRKN-34:

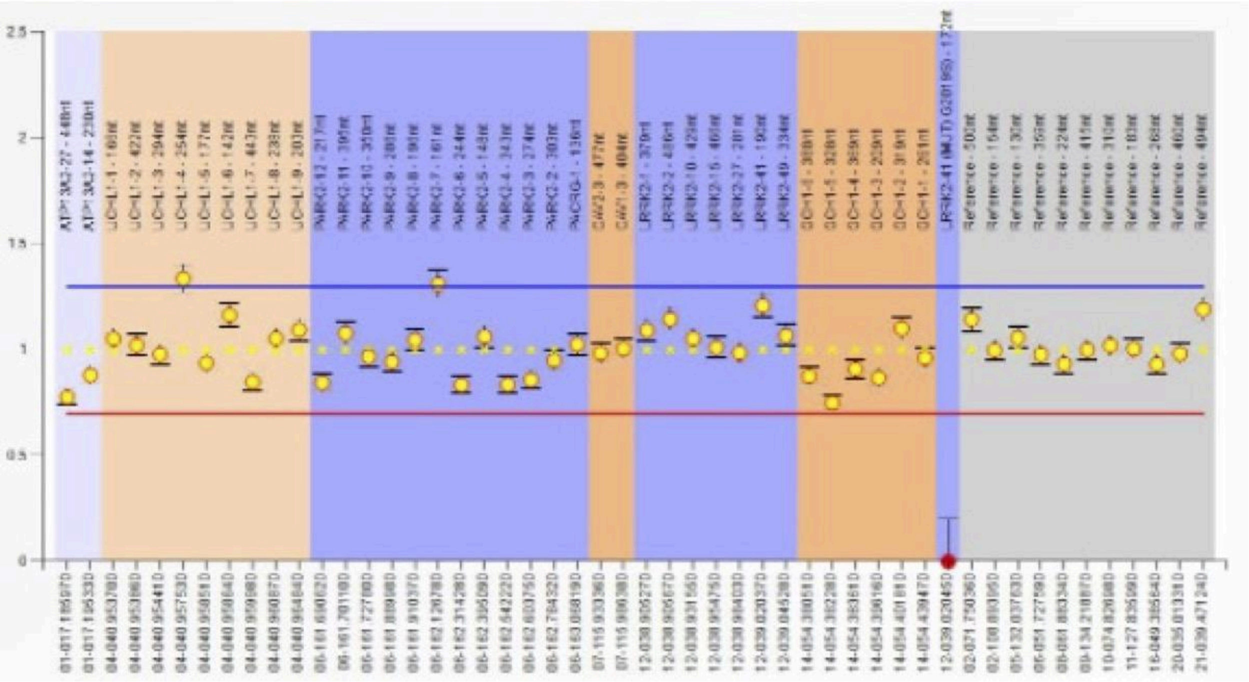

PRKN-35: Exon 6 duplication

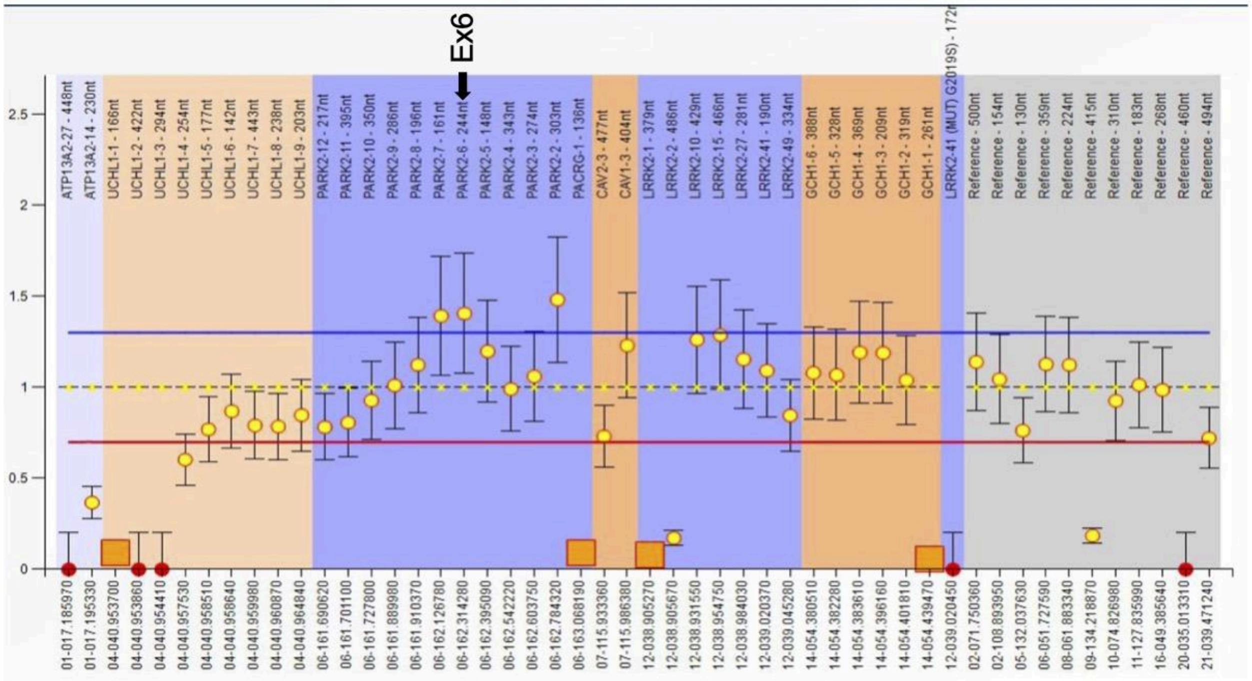


Ex6

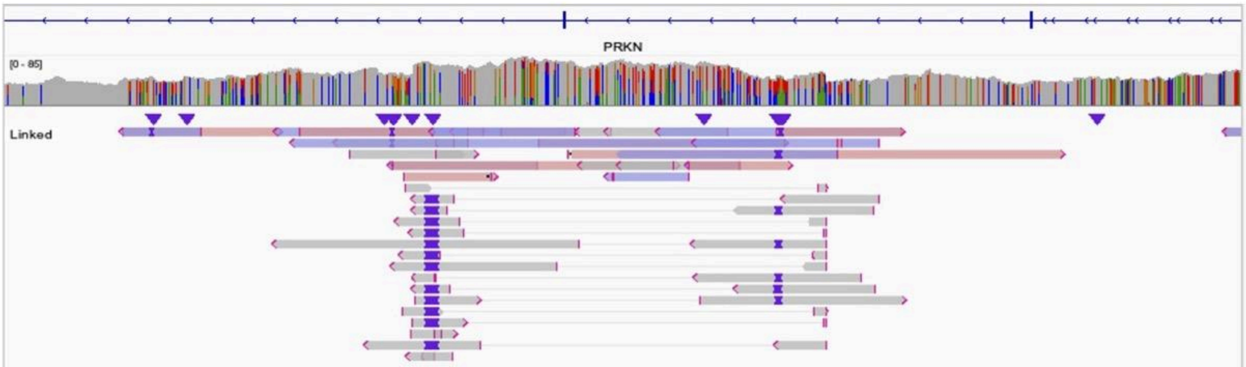

PRKN-36:

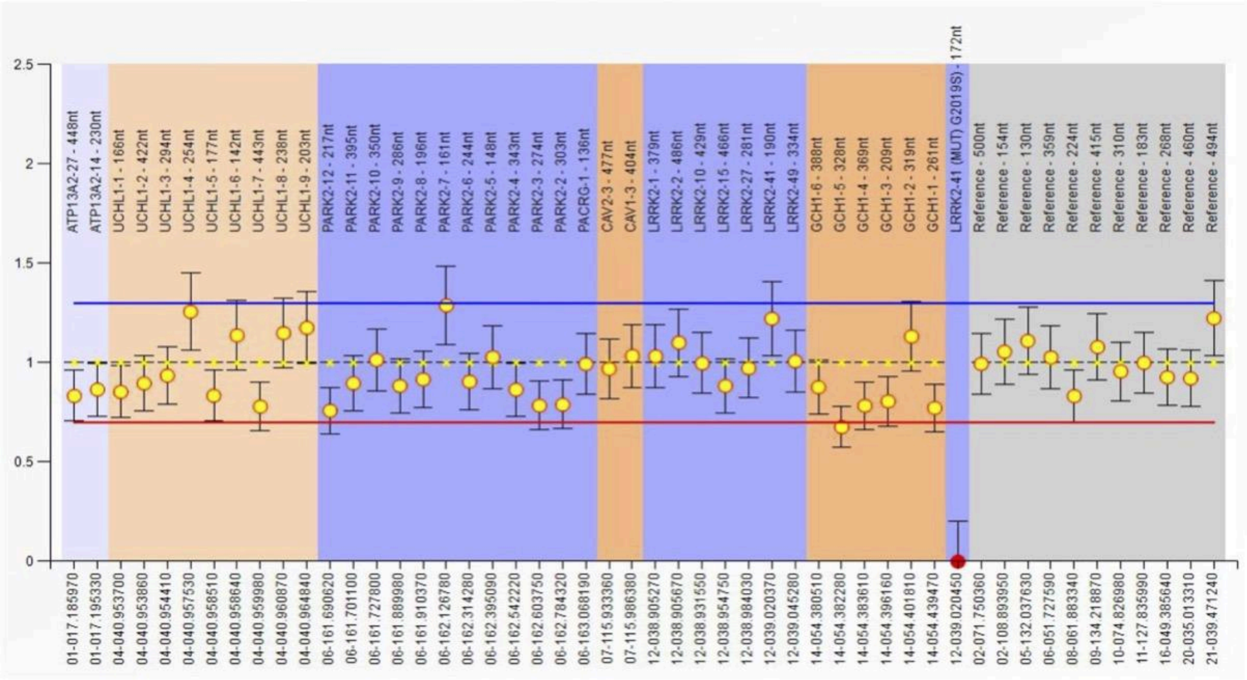


Ex; exon
