## Supplementary material for "The Utility of Long-Read Sequencing in Diagnosing Genetic Autosomal Recessive Parkinson’s Disease: a genetic screening study": eFigure3

### eFigure 3 Screenshot of IGV for DUP-NML-DUP/INV

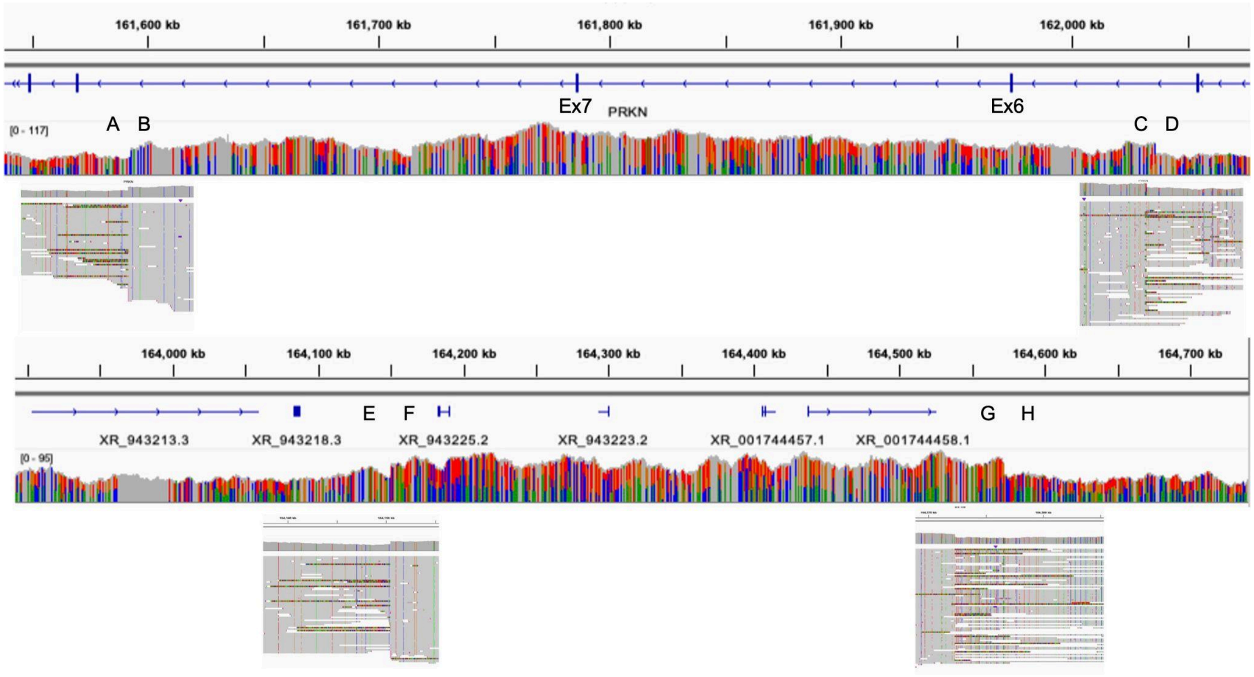


The upper panel showed the region including exon six and seven of *PRKN*. The lower panel showed the region 3 Mb away from the upper panel. The reads mapped at two breakpoints (A-B and C-D) of the upper panel were split but not aligned to each other. The split reads were aligned in the breakpoints in the lower panel (E-F and G-H).

IGV; integrative genome viewer, DUP; duplication, NML; normal, INV; inversion
