## Supplementary material for "The Utility of Long-Read Sequencing in Diagnosing Genetic Autosomal Recessive Parkinson’s Disease: a genetic screening study": eFigure4

**eFigure 4 PCR for junction of DUP-NML-DUP/INV**

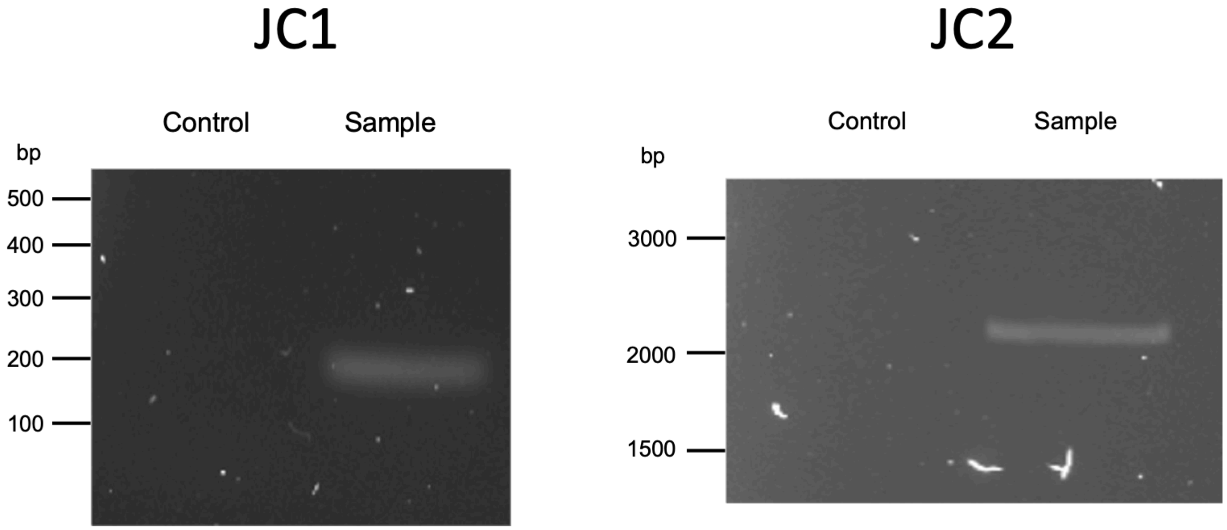


The expected length of the band is 208 bp in JC1 and 2166 bp in JC2. JC1 and JC2 correspond to JC1 and JC2 in Figure 3.
DUP; duplication, NML; normal, INV; inversion, JC; junction, bp; base pair
