## Supplementary material for "The Utility of Long-Read Sequencing in Diagnosing Genetic Autosomal Recessive Parkinson’s Disease: a genetic screening study": eFigure5

### eFigure 5 Correlation between the number of *PRKN* variants number and clinical phenotype

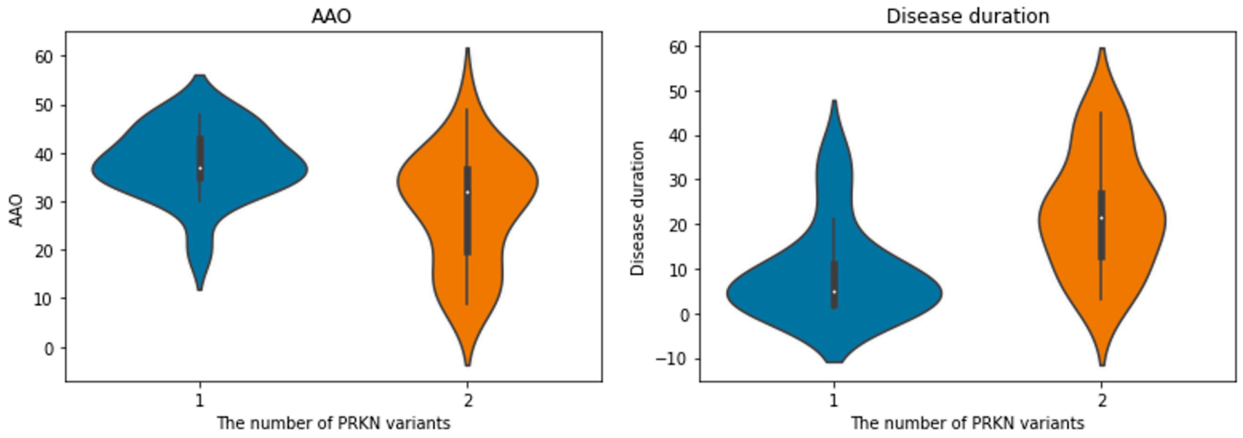


Correlation between the number of PRKN variants and the clinical phenotype with continuous value in which the p value was smaller than 0.05.
AAO; age at onset
